# An age-stratified mathematical model to inform optimal measles vaccination strategies

**DOI:** 10.1101/2025.04.01.25325066

**Authors:** Samiran Ghosh, Indrajit Ghosh, Siuli Mukhopadhyay

**Author notes:** Corresponding author *Email address:* (Siuli Mukhopadhyay). These authors contributed equally to this work (Samiran Ghosh, Indrajit Ghosh).

## Abstract

Measles remains a significant global public health threat despite the availability of an effective vaccine. WHO recommends the first measles vaccine dose at 9–12 months of age, balancing maternal antibody interference and immune system maturity. However, infants younger than 9 months remain highly vulnerable, especially in regions with high transmission and low herd immunity. This study introduces an age-stratified, multi-compartmental model of measles transmission that captures infections in unvaccinated infants by explicitly modelling maternal immunity decay and early infection risks. The model’s positivity is proven, and an analytical expression for the instantaneous replacement number is derived. For a non-age-stratified version, equilibrium points and their stability are analyzed. Using a bootstrap algorithm, critical parameters and age-specific transmission rates are identified by fitting the model to yearly incidence data from six measles-prevalent countries. The proposed model provides a framework for evaluating hypothetical vaccination scenarios, including different routine and supplementary dose schedules. Under the model assumptions, scenarios including MCV0 produced larger reductions in projected measles incidence than those without MCV0. The study estimates the relative number of cases averted and projects elimination timelines for several hypothetical scenarios, highlighting that combining routine and supplementary immunizations with alternative assumptions such as early measles vaccination may be highly effective.

## 1. Introduction

Measles, an extremely contagious viral disease, continues to pose a major global public health challenge despite the availability of a highly effective vaccine. Measles routine immunization (RI) activities as recommended by World Health Organization (WHO) consist of two doses of the measles vaccine, measles containing vaccine 1 (MCV1) and measles containing vaccine 2 (MCV2), respectively, at 9-12 months and 16-24 months of age, with at least 95% coverage [1].

In addition, several measles-endemic countries conduct supplementary immunization activities (SIAs) through which vaccines are administered to specific age groups regardless of their RI status [2]. Although vaccination has been successful in preventing an estimated 60 million deaths between 2000 and 2023, the disease is still prevalent in large parts of Africa and Southeast Asia. From the RI data, we note that in Southeast Asia, vaccination coverage has steadily increased to 91% for MCV1 and 85% for MCV2, however, several regions in those parts of the world are still recording frequent measles outbreaks. Thus, there is an urgent need to identify the factors responsible for these continued measles outbreaks and to investigate alternative measles vaccination strategies.

WHO recommends that the MCV1 be administered at 9 to 12 months of age, based on the risk of maternal antibody interference and the time of maturity of the infant’s immune system [3]. However, infants younger than 9 months remain vulnerable to measles [4], particularly in areas with high transmission rates and low herd immunity. A surveillance study suggested that 12% of measles cases reported to WHO were too young to be vaccinated [5]. This is of great concern as [6] notes a higher risk of developing severe symptoms in measles-infected infants of less than 9 months of age. Also, the proportion of measles cases in infants is likely to increase as more mothers have vaccine-induced immunity, resulting in their children having lower maternal antibody levels as compared to those of naturally immune mothers [4]. Recent studies suggest that vaccinating infants aged 6-9 months could bridge this vulnerability gap and contribute to better measles control [4]. However, higher protection and seroconversion were seen in children who received MCV1 at a higher age. The optimal age of vaccination of MCV1 particularly, is still an active area of research which depends on various factors such as the maturity of the immune system to respond to MCV, the age at which children are at risk of infection and the objectives of national immunization programmes.

A survey analysis based on case-based measles surveillance data from WHO headquarters, spanning 2011-2016, reveals a significant number of measles cases in the 6-8 month age group [7] (see Figure 1). Infants under one year of age are particularly vulnerable, as they are typically too young to receive immunization under most current vaccination guidelines and are no longer sufficiently protected by maternally transmitted antibodies. These maternal antibodies generally provide protection for an average of 3–6 months in infants born to naturally immune mothers, but this period is even shorter for infants of vaccinated mothers [8, 9, 10]. This evidence underscores the critical need to reduce measles cases among children younger than 9 months. In fact, countries like South Africa, Cambodia and Malaysia (Sabah state) have implemented MCV1 vaccination at 6 months whereas Sri Lanka and Colombia have administered SIA measles vaccination starting at 6 months age group [11, 12]. Although early vaccination offers a potential solution, its impact is limited by the reduced effectiveness of vaccines in younger age groups [13, 14]. This highlights the necessity of re-evaluating the balance between the benefits of early vaccination and the challenges posed by diminished vaccine effectiveness.

**Figure 1.**
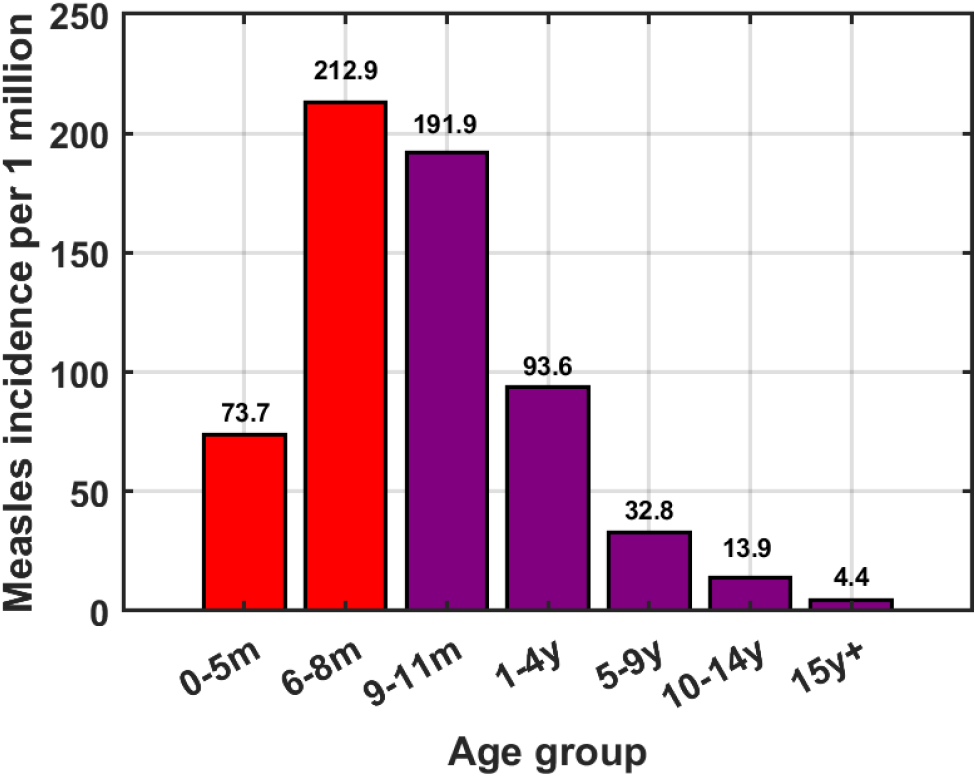
Average annual age-specific incidence of measles during 2011-2016 (adapted from [7]).

Age-stratified compartmental mathematical models have been widely used to study the dynamics of measles and evaluate the impact of vaccination strategies. Age stratification is particularly relevant for measles, as susceptibility, transmission, and immune responses vary significantly across age groups due to waning maternal antibodies and different susceptibilities [15]. By integrating demographic data, vaccination coverage, and disease parameters, these models provide a robust framework for identifying high-risk age groups and optimizing location-specific vaccination strategies [2, 16]. Several age-structured measles models have been previously employed to study vaccination scenarios. Verguet et al. [17] developed an age-stratified measles model called DynaMICE to compute the maximum allowable time between two consecutive SIAs for achieving measles control. They found that multiple SIAs at high coverage levels and regular intervals is a viable strategy to prevent measles outbreaks. Prada et al. [18] found that a single high-coverage SIA can effectively maintain measles elimination, with follow-up campaigns potentially requiring smaller investments in 4 African populations. Trentini at. al. [16] calibrated a compartmental model for 9 countries using historical serological data to estimate disease burden reduction and age-specific susceptibility. Another compartmental modelling study found that current vaccination policies are not sufficient to achieve and maintain measles elimination countries under study. They concluded that strategies targeting unvaccinated children before they enter primary school can remarkably enhance the fulfilment of WHO targets. Fu et al. [20] observed that high coverage of both RI doses and SIA is essential to achieve and maintain measles elimination goals. Winter et al. [21] concluded that achieving and maintaining the elimination of measles and rubella worldwide will require innovative vaccination strategies, technologies to address the inequities in routine coverage, and continued investment in surveillance and outbreak response. Auzenbergs et al. [2] found that to achieve high levels of vaccination coverage and meet targets for measles elimination in areas of high burden, SIAs must be strengthened and made more efficient, but routine two-dose coverage must also be improved over time. Recently, Goult et al. [22] used an age-stratified compartmental model to estimate the optimal age for infant measles vaccination. They found large heterogeneity in the optimal MCV1 ages, ranging 6–20 months of age. They showed that the optimal MCV1 age depends on the local epidemiology, with a lower optimal age predicted in populations having lower vaccination coverage or suffering from higher transmission.

The requirement of controlling measles in infants under 9 months of age remains an important problem in measles-endemic countries. In addition, very few of these models were calibrated according to the observed country-specific measles incidence data. The literature on examining the potential effect of vaccination of children in younger age groups (*<* 9 months) through mathematical models is sparse. Therefore, there is scope to study model-based projections of measles elimination for various scenarios, such as early routine MCV1 vaccination, additional doses of vaccine before 9 months of age along with SIAs at different frequencies combined with RI.

In this work, we propose a new age-stratified compartmental model of measles transmission to take into account measles infections in children who are too young to be vaccinated. The novelty of our model lies in the incorporation of maternal immunity loss and infection progression within the maternal immunity compartment, addressing a crucial dynamic that affects disease susceptibility over time in very young age groups. To achieve the elimination of measles by addressing the reduction in measles cases in the very young age group, we explore the following vaccination strategies primarily.

- Strategy 1: Routine MCV1, MCV2 with SIAs at different frequencies.
- Strategy 2: Early MCV1 (with reduced efficacy), routine MCV2 with or without SIAs at different frequencies.
- Strategy 3: Routine MCV1, MCV2 with SIAs at different frequencies and additional zero dose measles (MCV0) at 6-9 months once every three years.

The zero dose of the measles vaccine, known as the MCV0 dose or supplementary dose, is specifically recommended for infants aged 6 to 9 months in selected geographies where the risk of measles infection is particularly high (see page 37 in WHO: Measles and Rubella Strategic Framework 2024–2030). Although routine immunization schedules typically begin at 9 months of age with the MCV1 dose, the MCV0 dose acts as an early protective measure for vulnerable populations, especially during outbreaks or in settings with elevated exposure risk [23]. In strategy 3, we implement the zero-dose option at a single time point in a given year and not as part of RI activities.

In addition, we calibrate our proposed model to country-specific measles data for six countries where measles is prevalent, namely Nigeria, Philippines, Indonesia, Democratic Republic of the Congo (DR), India, and Pakistan. We use MCV1 and MCV2 coverages, SIA coverages, and demographic variables (population, birth rate, death rate) as inputs to the model. The primary contribution of this work is the development of a mathematical framework for comparing hypothetical vaccination scenarios. The proposed model enables quantitative assessment of the epidemiological impact of alternative assumptions such as early measles vaccination and other intervention strategies.

## 2. Model description

In this section, we propose an age-stratified model for studying the effects of early vaccination on measles transmission. While the age distribution of measles infections can shift to older cohorts in pre-elimination settings, fine age stratifications for children under 5 years (0-3, 3-6, 6-9, 9-12 months, and 1-year intervals up to 5 years) are employed in this study to capture key developmental and immunological changes, particularly the waning of maternal immunity and to evaluate the early infant vaccination strategies. Broader intervals (5-15 years and 15-80 years) are utilized to capture the remaining transmission dynamics within the older population. The population is divided into eight epidemiological compartments: maternally immune (*M* (*t*)), susceptible (*S*(*t*)), infected (*I*(*t*)), recovered (*R*(*t*)), vaccine failures (*F* (*t*)), successfully vaccinated with MCV1 (*V* ^1^(*t*)), successfully vaccinated with MCV2 (*V* ^2^(*t*)), and successfully vaccinated through SIA (*V* ^SIA^(*t*)), where *t* represents chronological time. Successfully vaccinated children are those who have received any dose of the measles vaccine and have developed immunity to measles.

We assume that children in class *M* are partially immune and that their immunity decays as they age. The subscript *i* denotes the compartments in a particular age group *i*. The youngest age group, 0 to 3 months, is represented by *i* = 1, and the index increases with age groups. The oldest age group, which includes individuals aged 15 to 80 years, is indicated by *i* = 10. Maternally immune children can become fully susceptible due to loss of maternal immunity. Susceptible individuals can either acquire infection or receive vaccination. Vaccination can lead to successful immunization or failure, in which case individuals remain susceptible and may require additional doses [16]. If infected, individuals move to the *I* compartment, where they contribute to disease transmission before moving to the *R* class, gaining long-term immunity. The flow diagram of the proposed model for a single age group, excluding births and deaths, is shown in Fig. 2. We denote this model as the primary model. Later, we introduce an alternative model to implement strategies 3 and 4.

**Figure 2.**
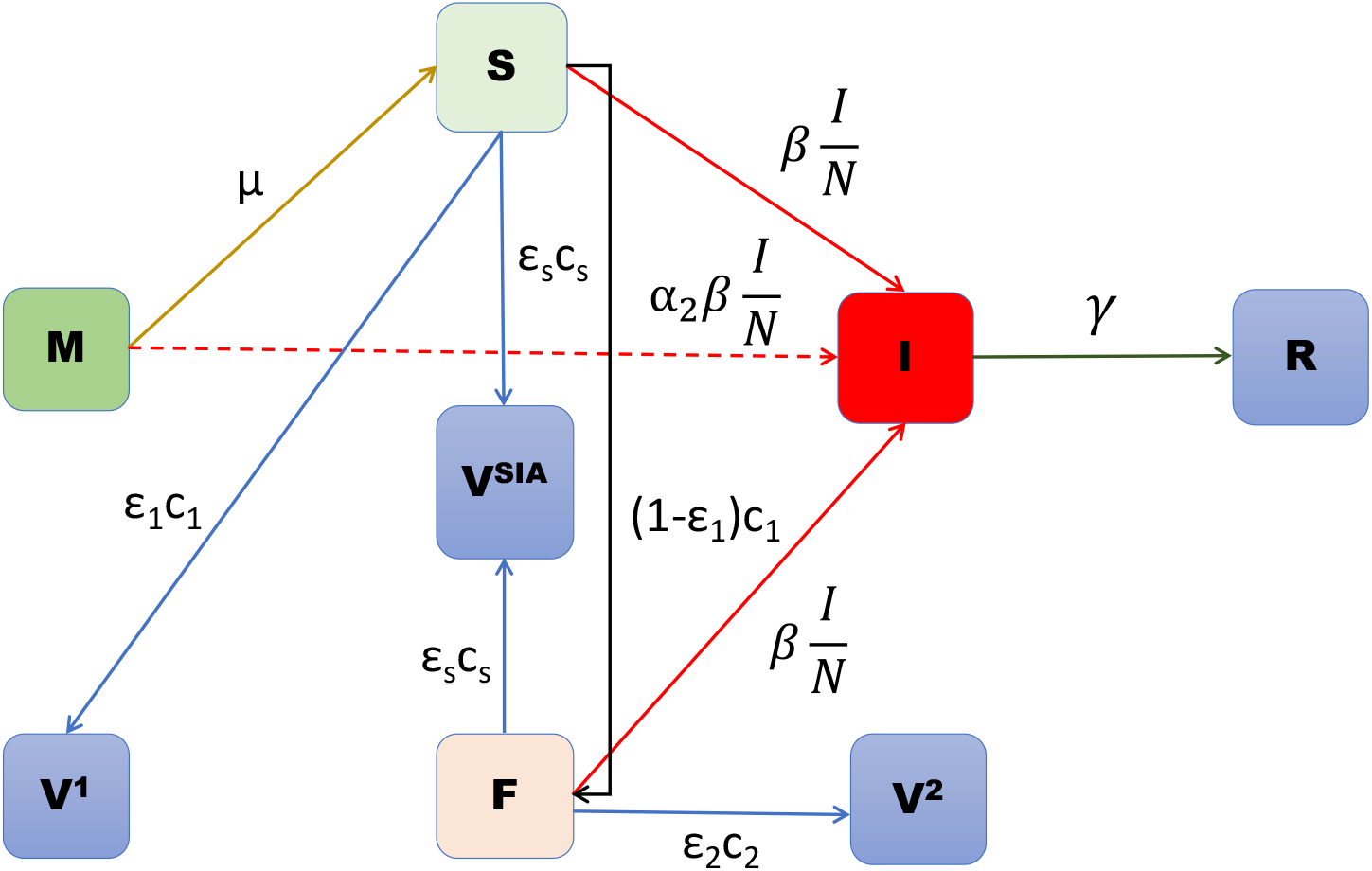
Flow diagram of the proposed model for a single age group without births and deaths. The dashed line from *M* to *I* represents the possibility of maternal immunity loss below 9 months of age.

To reiterate, the proposed model incorporates loss of maternal immunity and progression of infection in the maternal immunity compartment, allowing us to focus on the problem of susceptibility to measles over time in very young age groups. This addition is significant because it captures an important subgroup that could contribute to disease transmission and is usually ignored in the literature on measles modelling. A similar model was explored by [16], but unlike their approach, our model explicitly incorporates the interaction between infection risk in maternally immune individuals and the reduction in vaccine effectiveness due to early measles vaccination in children 6-9 months of age, offering a more comprehensive understanding of how these factors shape the dynamics of the measles epidemic.

The dynamics of disease transmission are described by the following system of differential equations:

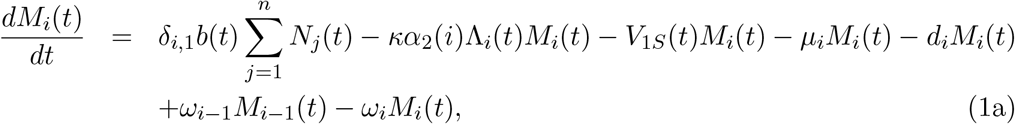

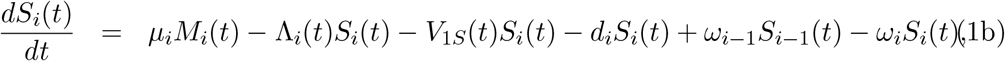

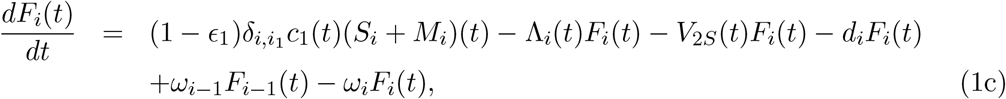

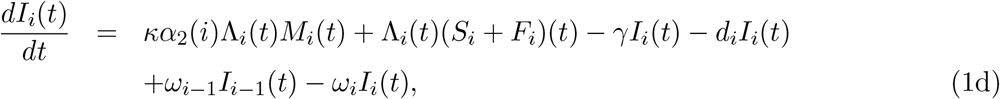

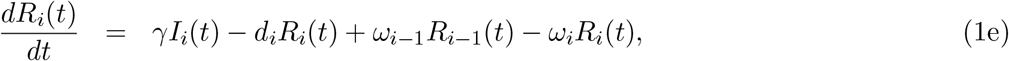

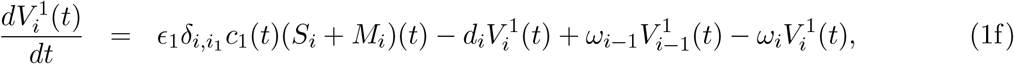

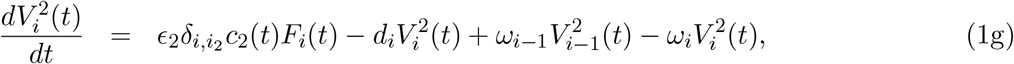

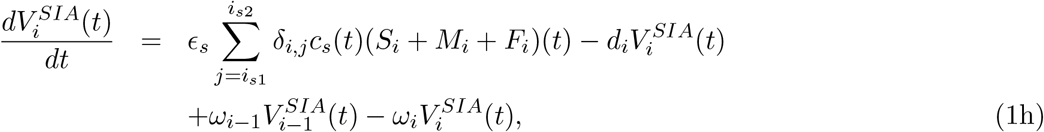

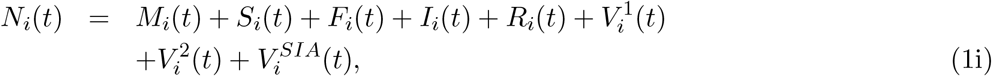

where

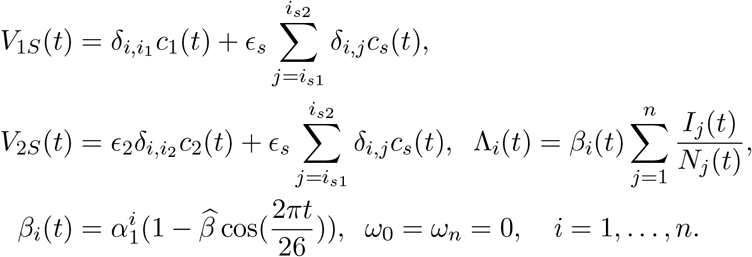

Here *n* represents the number of age groups. The birth and natural death rates in age group *i* are denoted by *b*_*i*_ and *d*_*i*_, respectively. The disease dynamics are governed by the recovery rate *γ* and the transfer rate *µ*_*i*_ from maternally immune individuals (*M*_*i*_) to susceptible individuals (*S*_*i*_) within each age group. The transmission dynamics are further characterized by *β*_*i*_(*t*), the age-specific transmission rate, which includes seasonal variations controlled by 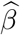, and 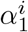, the age-specific rate of transmission. For simplicity, the transmission rates are assumed to be the same within the four age groups under 1 year 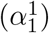, the same within the four age groups between 1 and 5 years 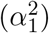, and the same within the two age groups between 5 and 15 years 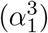, and the same for the 15+ years age group 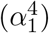. These rates are assumed to be different due to the variation in contact patterns among different age classes [24]. Additionally, *κα*_2_(*i*) represents an adjustment factor or transition probability that modifies the force of infection (Λ_*i*_(*t*)) for maternally immune individuals in age group *i*, reflecting the interaction between disease exposure and waning maternal immunity, lastly *δ* denotes the Kronecker delta function. Vaccination strategies are defined by the efficacy of the first dose (*ϵ*_1_), second dose (*ϵ*_2_), and SIAs (*ϵ*_*s*_), along with their respective coverage levels *c*_1_(*t*), *c*_2_(*t*), and *c*_*s*_(*t*). Unlike the efficacies of the first and second routine measles vaccine doses, reliable estimates of the effectiveness of SIA doses are not readily available in the literature. Therefore, as a conservative modelling assumption, we assume the efficacy of SIA vaccination to be lower than those of MCV1 and MCV2 (see Table 1). Here, *V*_1*S*_ and *V*_2*S*_ account for MCV1 coverage with SIA and MCV2 coverage with SIA, respectively. For MCV1, *ϵ*_1_*c*_1_ proportion of the susceptible population transition to *V* ^1^ compartment and remaining (1 − *ϵ*_1_)*c*_1_ proportion move to vaccine failure compartment. Similarly, for the second dose, only a proportion *ϵ*_2_*c*_2_ of individuals in class *F* can move to *V* ^2^. The term *ω*_*i*_ accounts for changes in the age-group distribution due to natural population growth in age group *i*.

**Table 1:** Description of the model parameters with corresponding values or ranges.

| Parameter | Description | Value/Range | Source |
| --- | --- | --- | --- |
| $n$ | Number of age groups | 10 | Assumed |
| $\alpha_1^i$ | Transmission rate in the age group $i$ | (0,5) | Estimated |
| $b$ | Birth rate | Country specific | World bank |
| $d_i$ | Natural death rate in the age group $i$ | Country specific | World bank |
| $\gamma$ | Recovery rate | 0.0714 | [16] |
| $\mu$ | Transfer rate from $M$ to $S$ | $1/(\text{length of third age group in bi-weeks}) = \frac{1}{6}$ | [25] |
| $\kappa$ | Modification factor | (0,1) | Estimated |
| $\alpha_2(i)$ | Relative susceptibility of class $M_i$ | $\alpha_2(1) = 0.2$, $\alpha_2(2) = 0.6$, $\alpha_2(3) = 1$ and $\alpha_2(i) = 0$ for $i = 4, \dots, n$ | [26] |
| $\epsilon_1$ | Efficacy of MCV1 | 0.95 | [17] |
| $\epsilon_2$ | Efficacy of MCV2 | 0.98 | [17] |
| $\epsilon_s$ | Efficacy of SIA | 0.70 | Assumed |
| $\hat{\beta}$ | Strength of seasonality | 0.05 | Assumed |
| $c_s(t)$ | SIA coverage | Country specific | [27] |
| $c_1(t)$ | MCV1 coverage | Country specific | WHO |
| $c_2(t)$ | MCV2 coverage | Country specific | WHO |
| $\omega_i$ | Rate of change of age-group from age group $i$ to $i + 1$, due to natural growth | $1/(\text{length of age groups in bi-weeks})$ | [25] |
| $\rho$ | Reporting rate | (0,1) | Estimated |

The targeted age groups for vaccination are defined by the parameters *i*_1_, *i*_2_, 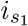, and 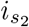. Here, *i*_1_ denotes the age group in which the first dose (MCV1) is administered, while *i*_2_ represents the age group receiving the second dose (MCV2). The SIAs are targeted at age groups ranging from 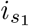 to 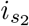. The flexibility of these parameters allows for the representation of various vaccination strategies. Mathematically, all parameters *i*_1_, *i*_2_, 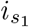, and 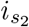 range from 1 to *n*. The model parameters and some descriptions are reported in Table 2.

**Table 2:** Table of estimated parameters for the six countries with mean and corresponding confidence intervals.

| Parameters<br>Countries | $\alpha_1^1$ | $\alpha_1^2$ | $\alpha_1^3$ | $\alpha_1^4$ | $\kappa$ | $\rho$ |
| --- | --- | --- | --- | --- | --- | --- |
| Nigeria | 2.588<br>(1.048 – 4.316) | 0.088<br>(0.001 – 0.322) | 0.15<br>(0.01 – 0.25) | 0.0134<br>(0.001 – 0.05) | 0.535<br>(0.066 – 1) | 0.0165<br>(0.004 – 0.055) |
| Philippines | 1.531<br>(1.3 – 2.4) | 0.092<br>(0.09 – 0.119) | 0.111<br>(0.08 – 0.25) | 0.008<br>(0.001 – 0.05) | 0.822<br>(0.385 – 1) | 0.0092<br>(0.009 – 0.0111) |
| Indonesia | 1.399<br>(1.118 – 2.1) | 0.102<br>(0.09 – 0.215) | 0.1004<br>(0.08 – 0.25) | 0.0078<br>(0.001 – 0.05) | 0.867<br>(0.454 – 1) | 0.01<br>(0.009 – 0.017) |
| DR Congo | 1.1128<br>(0.988 – 1.64) | 0.2355<br>(0.12 – 0.313) | 0.0749<br>(0.01 – 0.25) | 0.0204<br>(0.001 – 0.05) | 0.8914<br>(0.512 – 1) | 0.0031<br>(0.0025 – 0.005) |
| India | 0.8388<br>(0.731 – 0.954) | 0.5689<br>(0.378 – 0.75) | 0.0898<br>(0.01 – 0.25) | 0.0211<br>(0.001 – 0.05) | 0.9<br>(0.811 – 1) | 0.0045<br>(0.0033 – 0.0061) |
| Pakistan | 1.5288<br>(1.3 – 2.319) | 0.0951<br>(0.09 – 0.125) | 0.0839<br>(0.08 – 0.1356) | 0.0047<br>(0.001 – 0.0367) | 0.6727<br>(0.321 – 0.825) | 0.0091<br>(0.009 – 0.0108) |

## 3. Mathematical properties

### 3.1. Positivity and boundedness

Demonstrating the well-posedness of the model requires proving that it is both positive and bounded. Positivity guarantees that solutions remain within meaningful and realistic ranges, while boundedness ensures the solutions exhibit stable behavior. These attributes are crucial for establishing the model’s reliability and practical utility, instilling confidence in its analysis and predictive capabilities within the given context.

#### 3.1.1. Positivity

To prove the positivity of the system 1, we use the approach described in [28]. We set,

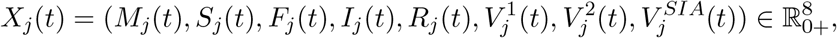

for *j* = 1, 2, · · ·, 10. Now for fixed *i* ∈ *{*1, 2, · · ·, 10*}*, we have the following:

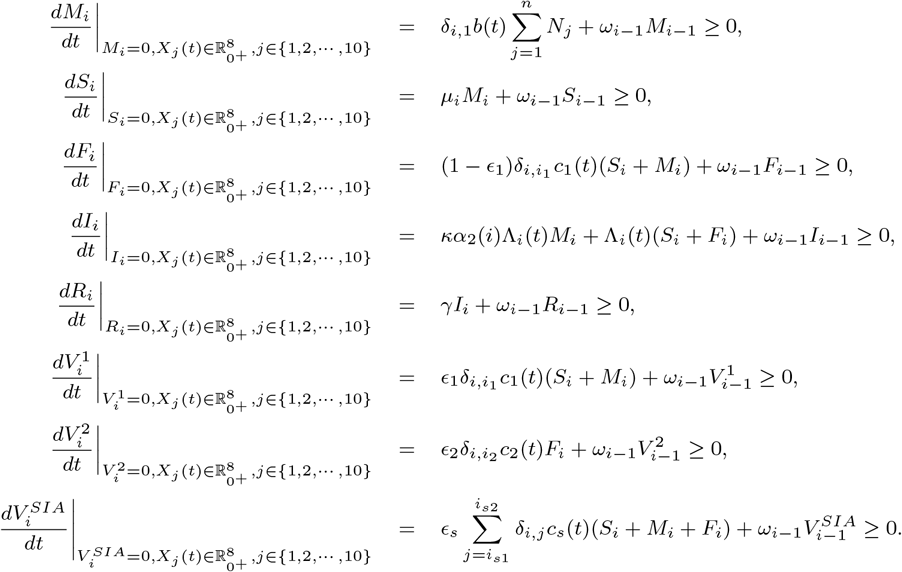

Hence, 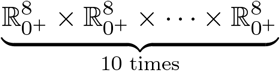 is invariant under the system (1). This completes the proof of positivity of the system (1).

#### 3.1.2. Boundedness

Adding all the equations in the system (1) we obtain:

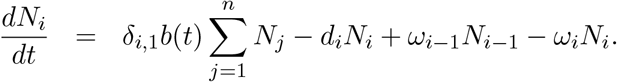

Suppose 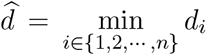, and 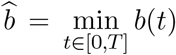, where *T* is the maximum time of observation. Then we can write

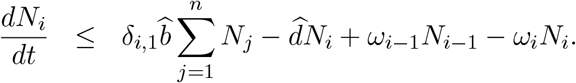

Suppose 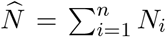, and *ω*_0_ = *ω*_*n*_ = 0. Summing *i* from 1 to *n* and assuming 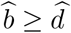 we obtain:

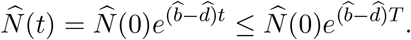

This completes the proof of positivity and boundedness of solution of the system 1.

### 3.2. Instantaneous replacement number

The instantaneous replacement number, ℛ_*t*_, is a fundamental metric for evaluating the transmissibility of an infectious disease and assessing the potential impact of control measures [29]. In this work we define instantaneous replacement number, ℛ_*t*_, as a threshold quantity such that at an instantaneous time *t*, if ℛ_*t*_ *<* 1 at time *t*, the total infected population in all age-groups will decay and if ℛ_*t*_ *>* 1 at time *t*, the total infected population in all age-groups will grow [29]. Since our model is age-stratified, we will find the instantaneous replacement number, ℛ_*t*_, for the whole infected population. Define, 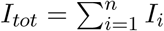. Taking sum over *i* in the equation (1d) from 1 to *n* and we obtain:

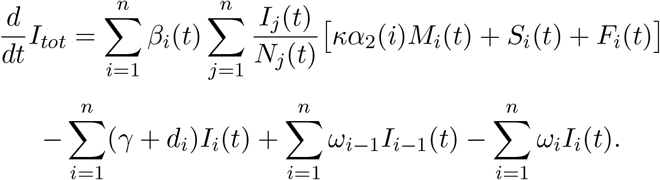

Since *ω*_0_ = *ω*_*n*_ = 0, we have 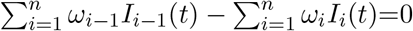. Consequently,

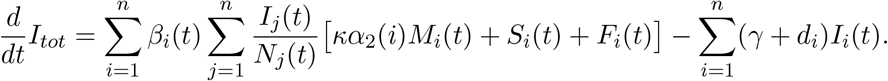

Thus the instantaneous replacement number, ℛ_*t*_ can be defined as follows:

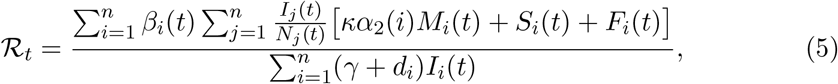

such that, at time *t*, if ℛ_*t*_ *<* 1 then 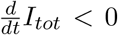 (i.e., the total infected population will not grow) and if ℛ_*t*_ *>* 1 then 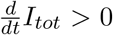 (i.e., the total infected population will grow).

Here, the numerator reflects the transmission potential normalized by the effective population at risk, while the denominator quantifies the overall rate at which infections are removed from the population, either by recovery or death. This characterization of ℛ_*t*_ will be utilized in the results section to analyze epidemic growth over the years across different countries.

Additional mathematical analysis on local and global stability was conducted for a simplified age-independent version of the model (1), see Appendix A for details.

In the following sections, using our proposed age-stratified model, we study the incidence and various strategies for the elimination of measles in six selected countries. A detailed description of the country-specific data that is needed as input to the model is provided. Estimation of age-specific transmission rates by calibrating the model with yearly measles incidence country-specific data is discussed. Using these calibration results, we analyze the sensitivity of model parameters on cumulative cases based on the partial rank correlation coefficients (PRCCs) method. The variation of the instantaneous replacement number estimated by the model over time is examined for each country. The effects of various vaccination strategies for the treatment of measles in very young age groups are studied and the relative number of averted cases for each country is estimated. We also explore the country-specific expected year for measles elimination under different strategies.

## 4. Data description

We consider data from six measles prevalent countries in Africa and South East Asian regions: Nigeria, Philippines, Indonesia, Democratic Republic of Congo (DR Congo), India and Pakistan [2]. The reported cases of measles from 2000 to 2023, the coverage of MCV1 and MCV2 for the six countries have been obtained from the WHO immunization dashboard https://immunizationdata.who.int/, while the coverage of SIA at the conutry-level for the period 2000 to 2020 has been taken from Auzenbergs et al. [2]. SIA administration ages for the countries Nigeria, Philippines, Indonesia, DR Congo and Pakistan are taken as 9-59 months, while for India, it is at age group of 9 months - 15 years. The initial population and initial age distribution of the countries are extracted from https://www.populationpyramid.net/. Immunization coverage of MCV1, MCV2 and SIA, along with WHO-reported cases for the six countries, is shown in Fig. 3. Among the six countries, India reports the lowest measles cases per million with at least 85% coverage for both MCV1 and MCV2. Note in the study period considered, the SIAs in India were mostly conducted at subnational level. However, since only country-level effective SIA coverage estimates were available in the public domain, we used these values as inputs in our national level model. The corresponding country-level coverages are given in Figure 3. We acknowledge that this is a limitation of the present study, as the country level coverages used may underestimate the impact of SIAs in settings where campaigns achieve high coverage within targeted regions.

**Figure 3.**
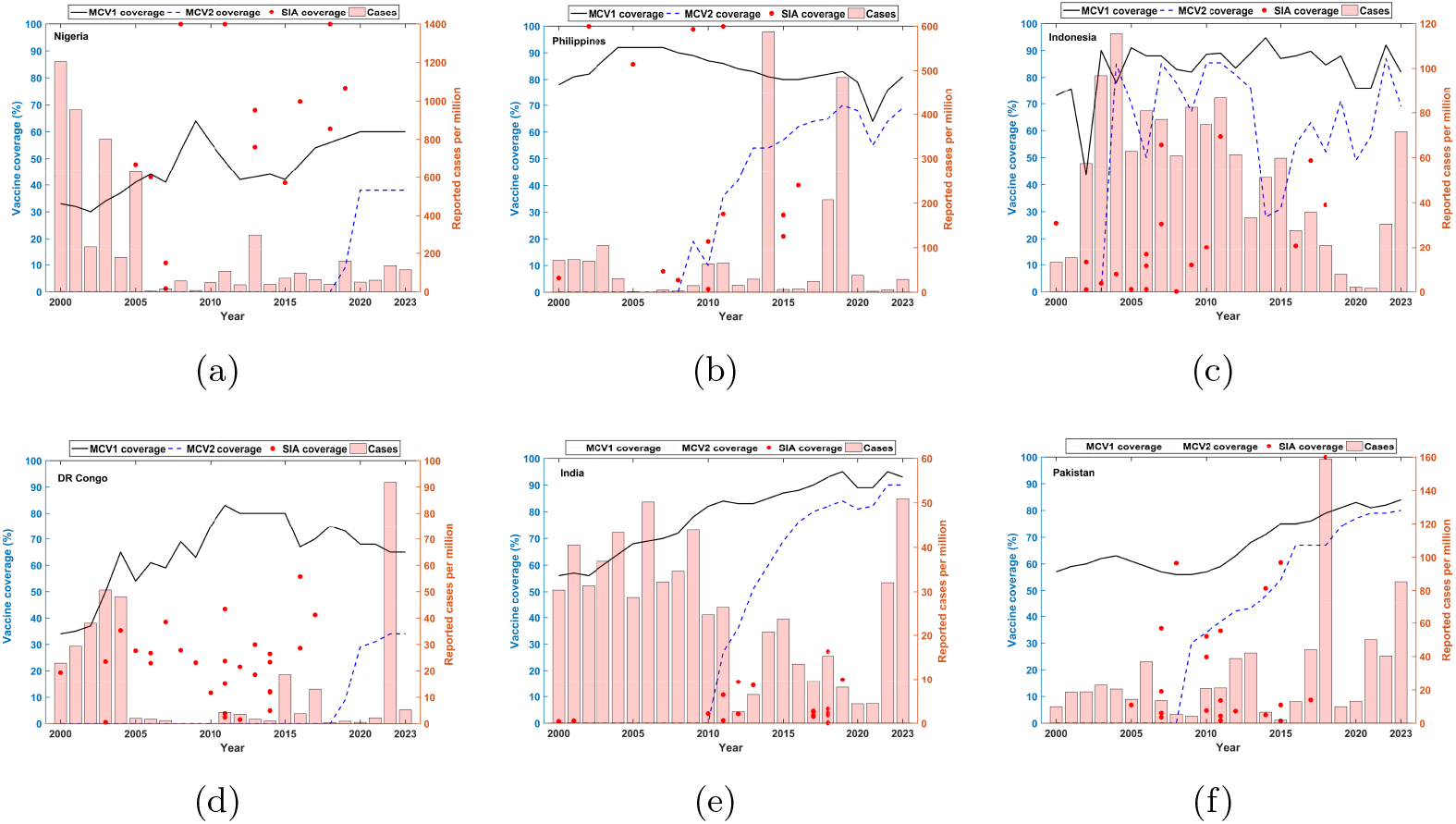
Immunization coverage for MCV1, MCV2 and SIA are depicted for - Nigeria, Philippines, Indonesia, DR Congo, India and Pakistan. Reported measles cases per million are also shown.

## 5. Model calibration

We calibrate the model (1) to yearly reported measles cases from the six countries of interest. Corresponding coverage data of MCV1, MCV2 and SIA for the six countries are given as inputs in the calibration process. The built-in optimization function *lsqcurvefit* in MATLAB (https://in.mathworks.com/products/matlab.html) was used to perform a non-linear least-squares fitting [30, 31]. We start the model simulations from 1970 for all the countries. The model parameters and the output for the infected compartment *I*(*t*) are defined in bi-weekly time units. We aggregated the bi-weekly incidence values over each year to obtain yearly totals. The initial conditions for DR Congo are reported in Table For initial conditions for other countries, please see the GitHub code available at https://github.com/nsamiran/Age-stratified-model.git. We assume that the initial susceptible population is 20% of the total population in the age group 9 months - 15 years in 1970. The total population and age-specific population for each country in 1970 were obtained from the population pyramid website (https://www.populationpyramid.net/india/1970/). Further, we assume that the initial age-wise susceptible is proportional to the number of infections in corre-sponding age groups as obtained from the IHME website https://www.healthdata.org/research-analysis/gbd-publications. In addition, we utilize the parametric bootstrapping technique to quantify the uncertainty of the parameters and obtain the 95% confidence intervals (CIs) of the estimated parameters [32, 33]. We generate 1000 bootstrap samples from the best-fitted model [32, 34] assuming a normal error structure. The mean and variance of the normal error structure are a single data point from the best fit curve and 0.2 *×* the same data point, respectively. For instance, if the best fitted curve is (*ŷ*_1_, *ŷ*_2_, …, *ŷ*_*n*_), then we obtain bootstrap samples by taking samples from normal distribution with mean *ŷ*_*i*_ and variance 0.2 *× ŷ*_*i*_. Instead of using a Poisson distribution to generate the bootstrap samples we use the asymptotic normal distribution. We chose the scale factor as 0.2 in the variance by a trial and error method for obtaining shorter confidence intervals for the parameters.

For the 1000 bootstrap samples/reconstructed time series data, the model was fitted using nonlinear least squares. The 1000 sets of parameters obtained from these bootstrapped time series are used to characterize the empirical distribution and the corresponding 95% CIs of the parameters. Consider the model (1) to be of the form

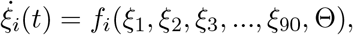

where 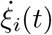 denotes the rate of change in the state variables *ξ*_*i*_(*t*) where *i* = 1, …, 90 and Θ is the set of model parameters. The steps for performing the model calibration are given below for a specific country:

- Derive the parameter estimates through least-square fitting using time series data *Y* = (*y*_1_, *y*_2_, *y*_3_, …, *y*_*n*_) to obtain the best fitted model.
- Generate bootstrap replications of the obtained best fitted model assuming a normal error structure.
- Re-estimate the parameters by fitting the model to the 1000 simulated time series data.
- Construct empirical distribution and confidence intervals of the estimated parameters.

A schematic flow diagram of the model calibration process in displayed in Fig. 4. The parameter estimates from the first step are reported in Table C.5 for all six countries. We assume that only *ρ* proportion of cases are reported for the countries. For simplicity and to avoid introducing additional complexity and identifiability issues in parameter estimation, we have assumed a constant reporting fraction in the present study. This assumption is also reasonable when the analysis is conducted at the country level. In a given period, outbreaks may occur in some states or regions while others may not experience outbreaks; consequently, the reporting fraction can vary across states. When aggregated at the national level, these regional variations can be reasonably approximated by a fixed average reporting fraction for the country. Therefore, we fit the model-generated infections multiplied by the reporting *ρ* with the actual reported cases to WHO. Using this method, we estimate six parameters: 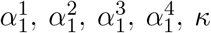 and *ρ*, for each country. The model fit is displayed in Fig. 5 for the six countries. These estimated parameter values presented in Table 2 are used in the numerical simulations presented in the following section.

**Figure 4.**
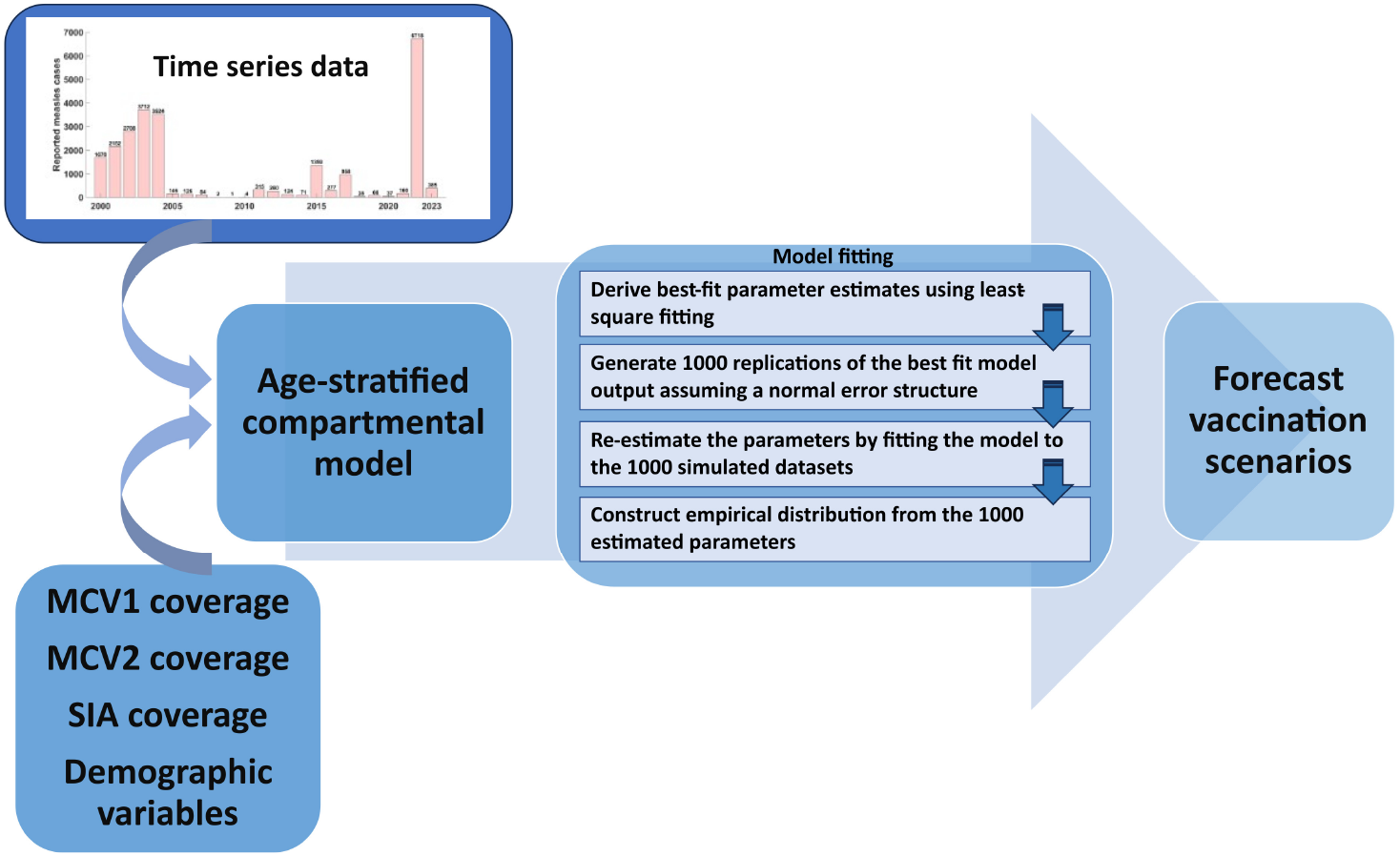
Flow diagram of the model calibration and forecast scenarios.

**Figure 5.**
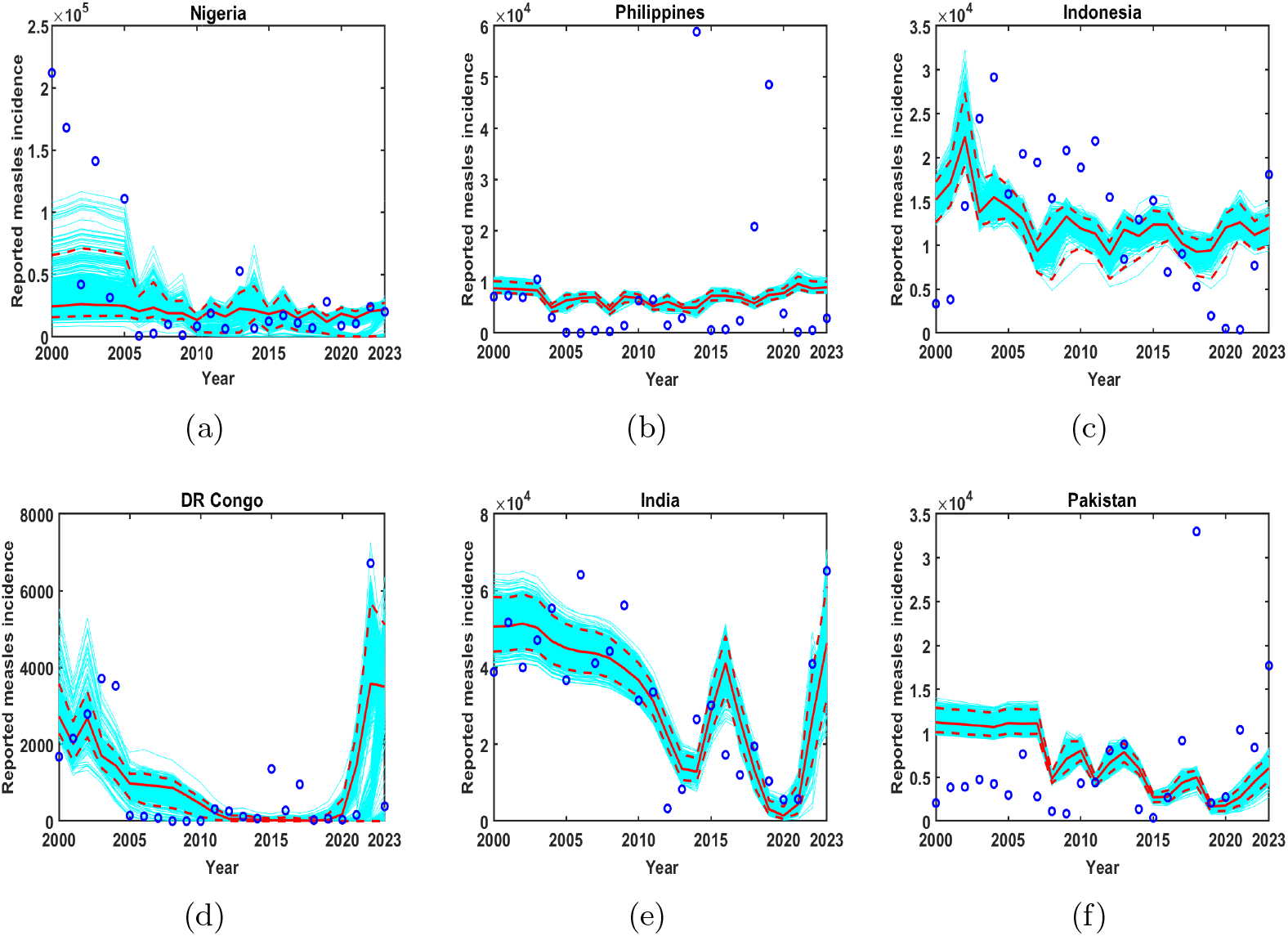
Fitting the model (1) to WHO reported measles cases in the countries Nigeria, Philippines, Indonesia, DR Congo, India and Pakistan respectively. The blue circles represent observed data, solid red line indicate mean model fit and the dotted red lines represent 95% CI.

## 6. Numerical results

In this section we perform numerical simulations based on the proposed age stratified model (1).

### 6.1. Sensitivity analysis

Global sensitivity analysis allows us to identify influential parameters while studying disease transmission. Here we use the PRCC method as discussed in Marino et al. [35].

The input parameters for the sensitivity analysis are: transmission rates for the different age groups 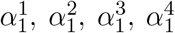; modification parameter for transmission rates of the maternal immune class, *κ*; respective coverage and efficacies of MCV1 and MCV2, *c*_1_, *c*_2_, *ϵ*_1_, *ϵ*_2_. The model (1) is simulated for a general hypothetical framework considering the effects of only routine doses of MCV1 and MCV2 vaccinations. The initial population size is taken to be 1 million. All parameters are varied in the range (0,1) except *ϵ*_1_ and *ϵ*_2_, these are, respectively, varied between (0,0.95) and (0,0.98). The response variable is the aggregate incidence of measles over 30 years 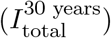. This global sensitivity analysis quantifies the impact of the parameters on the response variable. Latin Hypercube Sampling (LHS) is used in generating 5000 samples of each parameter from the corresponding ranges. The PRCCs are then calculated using the MATLAB functions provided in [35]. The results are shown in Fig. 6.

**Figure 6.**
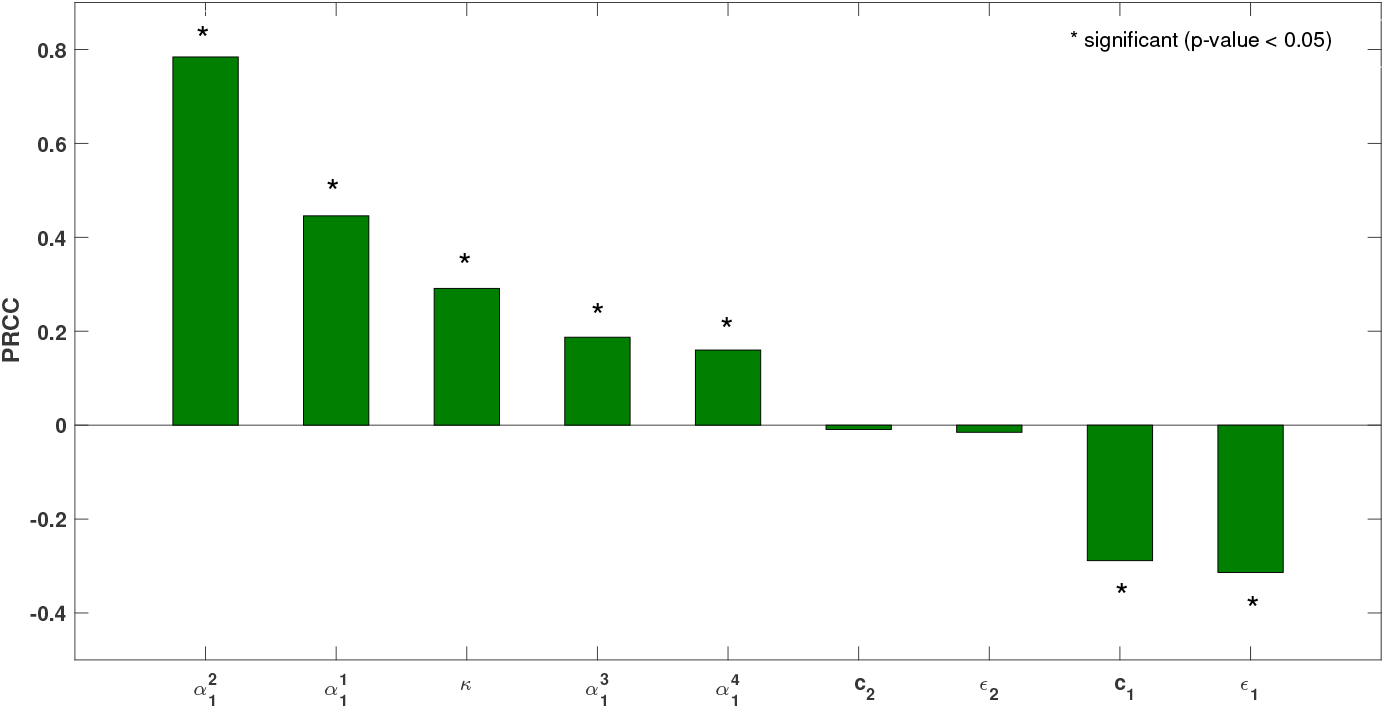
Sensitivity of model parameters with respect to aggregated measles incidence over 30 years. Significant PRCCs are indicated as **\*** (p-value *<* 0.05). Fixed parameters are taken as in Table 1.

It is observed that 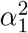, that is, the transmission rate for age groups 1-5 years is the most positively correlated parameter with the response variable 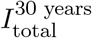, followed in decreasing order by 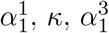 and 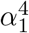. On the other hand, *ϵ*_1_ is the parameter most negatively correlated with 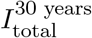, followed by *c*_1_. These results imply that while an increase in age group-wise transmission rates significantly (at level 5%) increases total measles cases in the next 30 years, an increase in coverage and efficacy of MCV1 vaccination significantly decreases measles cases in the next 30 years.

### 6.2. Estimates of instantaneous replacement number

The instantaneous replacement number is a key metric in assessing the transmissibility of an infectious disease, representing the average number of secondary infections generated by an infected individual in a population made up of both susceptible and non-susceptible hosts. Using parameter estimates from Table 2 and fixed parameter values from Table 1 in Equation (5), we plot the estimated ℛ_*t*_ for the six countries shown in Fig. 7. During the period 2020 to 2023, we observe that the ℛ_*t*_ values remain within the range (0.85, 1.15) with variations between countries, reflecting differences in vaccination coverage, demographic variables, and measles surveillance systems. A similar trend was observed in ℛ_*t*_ over the years in [36], with oscillations around the endemic situation.

**Figure 7.**
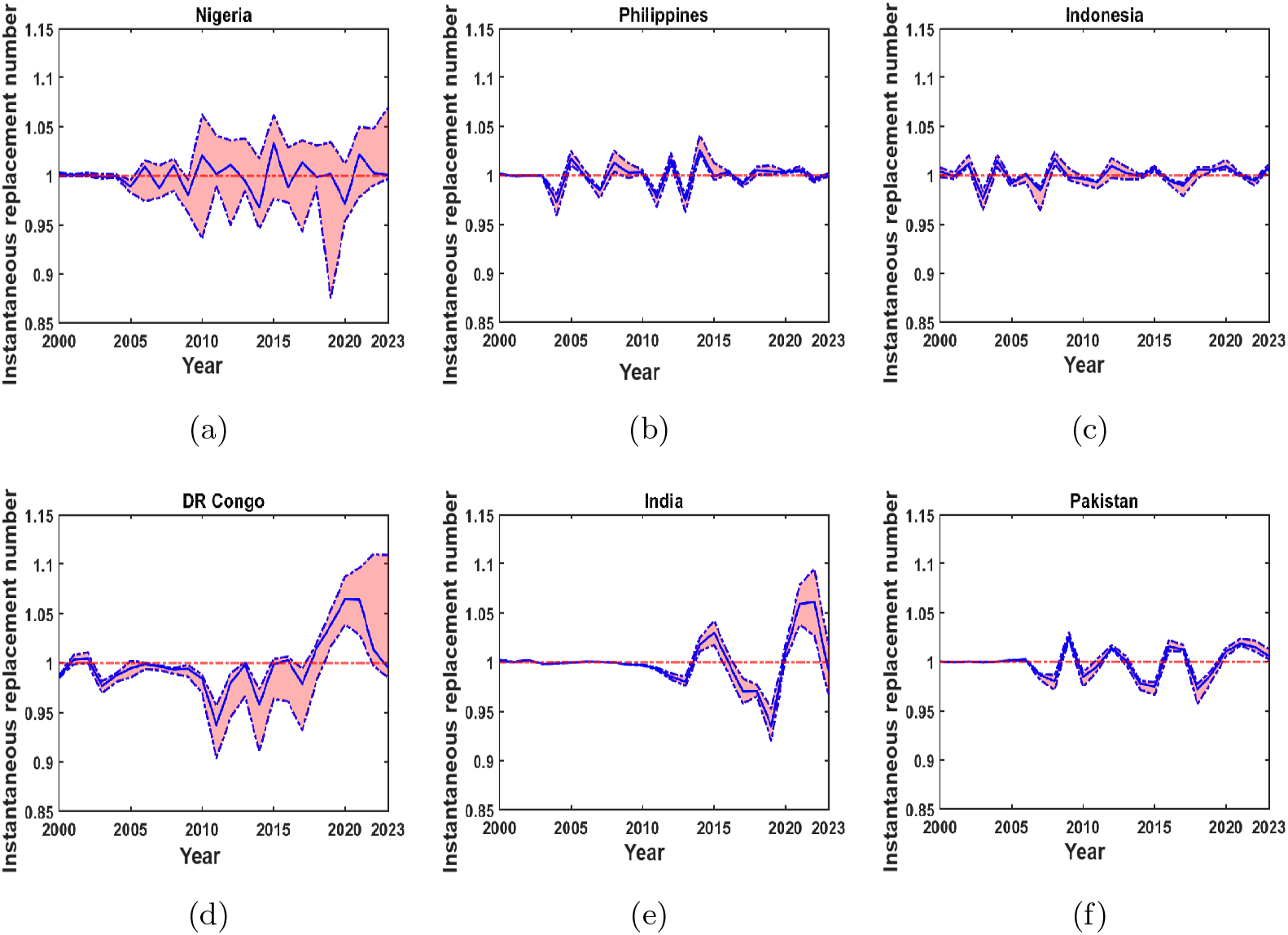
Plot of the estimated instantaneous replacement number over time from 2000 to 2023. The solid blue line represents the mean estimates, while the dashed blue lines indicate the 95% confidence interval. The horizontal red dashed line corresponds to ℛ_*t*_ = 1.

The slope of ℛ_*t*_ is positive during 2022 − 2023 in Philippines and Indonesia, indicating the possibility of future outbreaks unless remedial measures are taken. The mean values of ℛ_*t*_ are *>* 1 in multiple countries reflecting the need for more intense vaccination campaigns. For Nigeria and DR Congo, the 95% CIs of ℛ_*t*_ are wider than other countries, indicating the possibility of higher values of *R*_*e*_. During 2021-2022, the values of *R*_*t*_ for DR Congo, Pakistan, Nigeria, and India are seen to be (*>* 1), possibly indicating the effect of the pandemic on vaccination campaigns.

Differences in estimates of *R*_*e*_ over time and space reveal the varying effect of interventions in each country in any given year, while highlighting the need to tailor vaccination strategies to achieve measles elimination and mitigate measles transmission according to the epidemiological setting.

### 6.3. Scenario analysis

To assess the impact of vaccination interventions on measles control and elimination, we consider 3 strategies. Further, under each broad strategy, we study different scenarios (see Table 3). The early vaccination scenarios are motivated by some countries like South Africa, Cambodia, Sri Lanka, Colombia and Malaysia (Sabah state). South Africa, Cambodia and Malaysia (Sabah state) have implemented MCV1 vaccination at 6 months whereas Sri Lanka and Colombia have administered SIA measles vaccination starting at 6 months age group [11, 12]. We start with a baseline scenario that consists only of RI activities. RI with the standard timing of vaccination plus SIAs (with varying frequencies over years) form scenarios 1-3. Preponement of MCV1 to 6-9 months and MCV2 at standard timing with and without SIAs at different frequencies form scenarios 4-7. In scenarios 4-7, it is also assumed that MCV1 preponing induces 25% a reduction in vaccine effectiveness [37]. In scenarios 8 - 11 which are under strategy 3, we have RI combined with measles zero-dose (MCV0) at 6-9 months once every three years, and SIAs at different frequencies. To recap, Scenarios 1-3 represent *Strategy 1*, Scenarios 4-7 represent *Strategy 2*, and Scenarios 8-11 represent *Strategy 3*. For children over 9 months, the SIA coverages vary by country: 35% in Nigeria, the Philippines, Indonesia, and DR Congo; 16.5% in India; and 80% in Pakistan, which are based on the range of the last SIA coverages observed during the training period for each country [2]. We note that the SIA coverage values used for India represent country-level effective coverage estimates, whereas SIAs during the study period were predominantly conducted at the subnational level. Since subnational SIA coverage data were not publicly available, we used country-level effective coverage estimates computed following the DynaMICE methodology [17]. Although this may underestimate the impact of SIAs in targeted regions, it is appropriate for the country-level comparative analyses of broad vaccination strategies considered in this study. In Scenarios 8 - 11 with MCV0, we have considered the MCV0 coverage to be 28% for children aged 6 to 9 months in all countries. Specifically, for India in scenarios 8-11, 6-9 months, children receive doses with 28% coverage and over 9 months, children receive SIA doses with 16. 5% coverage.

**Table 3:**
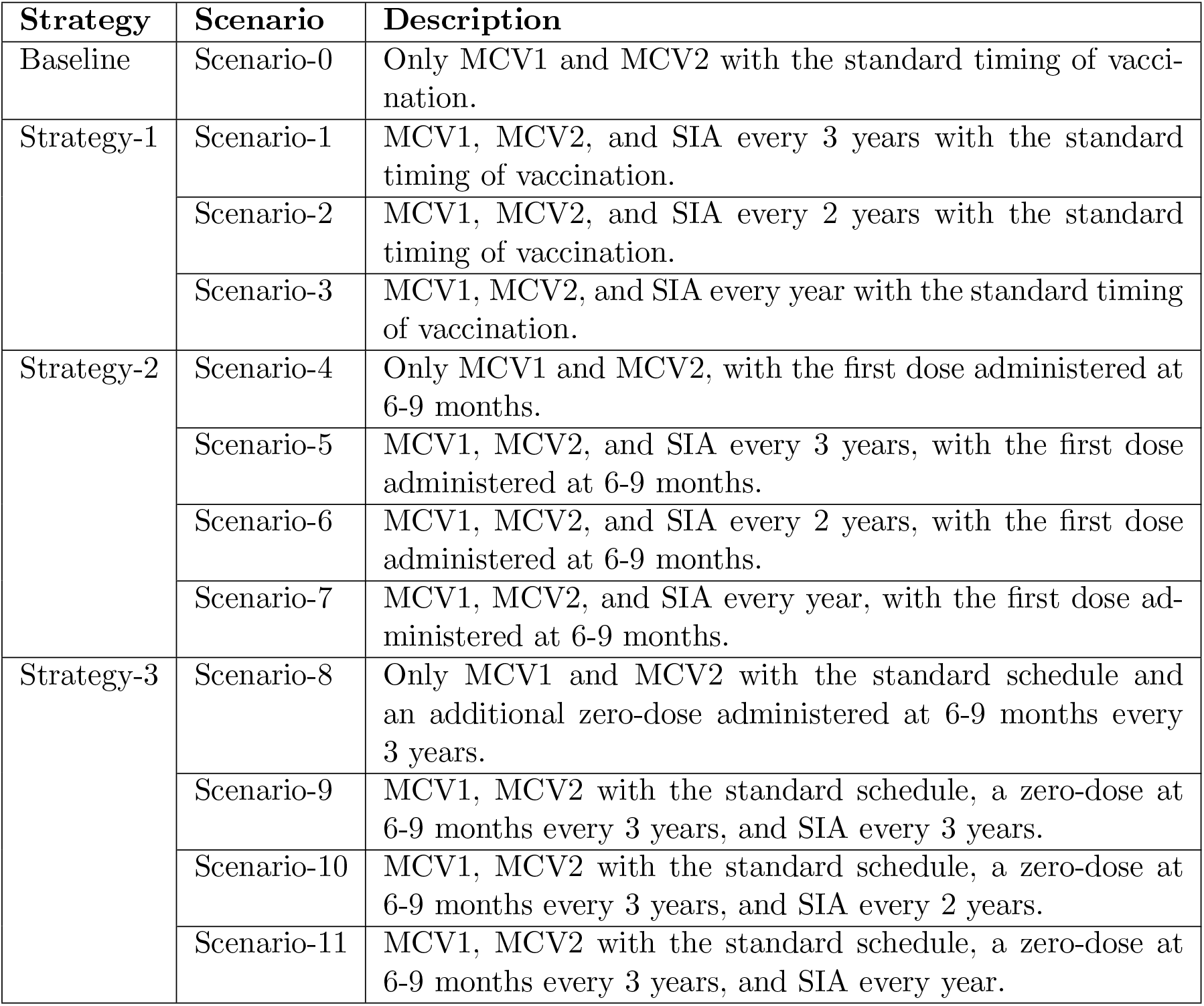
Summary of vaccination scenarios considered for analysis

| Strategy | Scenario | Description |
| --- | --- | --- |
| Baseline | Scenario-0 | Only MCV1 and MCV2 with the standard timing of vaccination. |
| Strategy-1 | Scenario-1 | MCV1, MCV2, and SIA every 3 years with the standard timing of vaccination. |
|  | Scenario-2 | MCV1, MCV2, and SIA every 2 years with the standard timing of vaccination. |
|  | Scenario-3 | MCV1, MCV2, and SIA every year with the standard timing of vaccination. |
| Strategy-2 | Scenario-4 | Only MCV1 and MCV2, with the first dose administered at 6-9 months. |
|  | Scenario-5 | MCV1, MCV2, and SIA every 3 years, with the first dose administered at 6-9 months. |
|  | Scenario-6 | MCV1, MCV2, and SIA every 2 years, with the first dose administered at 6-9 months. |
|  | Scenario-7 | MCV1, MCV2, and SIA every year, with the first dose administered at 6-9 months. |
| Strategy-3 | Scenario-8 | Only MCV1 and MCV2 with the standard schedule and an additional zero-dose administered at 6-9 months every 3 years. |
|  | Scenario-9 | MCV1, MCV2 with the standard schedule, a zero-dose at 6-9 months every 3 years, and SIA every 3 years. |
|  | Scenario-10 | MCV1, MCV2 with the standard schedule, a zero-dose at 6-9 months every 3 years, and SIA every 2 years. |
|  | Scenario-11 | MCV1, MCV2 with the standard schedule, a zero-dose at 6-9 months every 3 years, and SIA every year. |

We use parameter values from Tables 1 and 2 to generate the infections from the model (1) for the period 2024 to 2050. The routine MCV1 and MCV2 coverages in the years 2024 to 2050 are assumed to remain the same as of 2023 in a given country. Thus, we do not consider intensified vaccine coverages in the forecasting period. Furthermore, SIA administration ages for the countries Nigeria, Philippines, Indonesia, DR Congo, and Pakistan are taken as 9-59 months, whereas for India it is 9 months - 15 years age group [2]. Note the age cohort taken matches with the last SIA conducted in each country. The birth and death rates in the forecasting period are assumed to follow a linear trend similar to that in the calibration period.

For implementing the zero dose measles (MCV0) in strategy 3, we propose an alternative model (1) with added equations to incorporate the additional dose. Individuals who successfully acquire immunity following MCV0 vaccination are assigned to the (*V* ^0^) compartment. In practice, infants who receive MCV0 continue to follow the routine immunization schedule and are therefore eligible to receive both MCV1 and MCV2 at the recommended ages. For model simplicity, infants in whom MCV0 fails to induce immunity are assumed to remain in the (*M*) compartment, after which they enter the routine vaccination pathway according to the standard immunization schedule. Since MCV0 failure does not confer vaccine-induced immunity, this assumption preserves the intended vaccination sequence while avoiding the complexity of introducing additional compartments, and it does not alter the underlying transmission dynamics. The additional differential equation for *V* ^0^ and the modified equations for compartments, *M* and *V* ^1^, are given below.

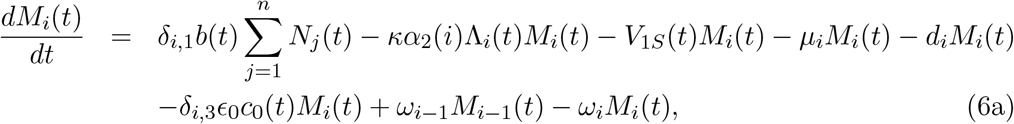

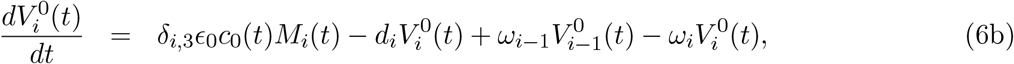

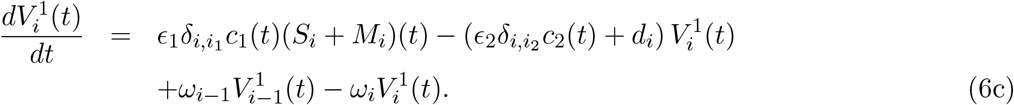

The efficacy of zero dose is indicated by *ϵ*_0_ with the corresponding coverage level *c*_0_(*t*) = 0.28. For zero doses, we use efficacy *ϵ*_0_ = 0.70 [37] same as *ϵ*_*s*_. To accommodate the additional compartment of *V* ^0^, in the flow diagram in Fig. 2, we should add the compartment *V* ^0^ that receives the inflow of individuals from the M compartment and is connected by an outward arrow to *V* ^1^.

The country-specific forecasted cases in the period 2024-2050 are shown in Figs. B.12 – B.17. For Nigeria, we note that the forecasted measles counts show a steady increase in case counts if Strategy 1 is followed. Preponement of MCV1 with and without SIAs forming Strategy 2 fails to stop the increase in the forecasted case counts. However, the use of additional doses of MCV0 under Strategy 3 is more effective in curbing the increase in the case counts though there are still 0.25 *×* 10^5^ reported cases in 2050. In Philippines, Strategy 1 is not helpful; however, Strategy 2 is able to decrease the case counts compared to strategy 1. As before, Strategy 3 brings down the reported measles cases. For Indonesia, we see a steep decrease in case counts with conventional Strategy 1. The cases are further reduced with the second Strategy. The addition of MCV0 to vaccination campaigns proves to be very successful, showing the possibility of elimination by 2030 with no resurgence. The partial success of the second strategy in both the Philippines and Indonesia may indicate higher infections in children younger than 9 months in these two countries. From the literature, we noted a recent outbreak in Indonesia where 10% of the cases were found in children less than 9 months [38]. Also, Domai et al. [39] reported the interquartile age range of measles infection in the Philippines to be between 7-28 months. Scenarios 10 and 11 are able to bring down the cases of measles to zero without further resurgence in Indonesia by 2030, while for the Philippines, it takes much longer. The forecasted incidence of measles in DR Congo shows a steep upward trend under RI activities. Adding SIAs to RI under Strategy 1, is not very effective in reducing reported cases unless the frequency of SIAs is high. Strategy 2 performs better as compared to Strategy 1, particularly with more frequent SIAs. The additional MCV0 combined with frequent SIAs shows that the cases drop to zero by 2030 with no further resurgence in DR Congo. The high coverages of MCV1 and MCV2 reported in India show continued RI activities at these coverages, and combining them with SIAs is effective in reducing the burden of measles. Strategy 2 shows a small improvement over Strategy 1 in the forecasted measles cases. The addition of MCV0 to the vaccination schedules and increasing the frequency of SIAs shows a drop in the measles cases before 2030 with no further resurgence till 2050. In-sample measles cases in Pakistan are on average much lower than other countries except India, this may be due to low reporting rates as noted previously by [40, 32]. This large under-reporting of cases is suspected to produce lower forecasted measles case values. For Pakistan, almost all strategies seem to be effective in lowering future cases, showing an early decrease in measles before 2030 with no further resurgence. Also, RI combined with high-frequency SIAs (scenario 7) gives slightly better results than those with MCV0. From these results, it is evident that SIA, pre-exposure of MCV1 to 6-9 months and adding a measles zero-dose have different rates of beneficial effects on reducing the measles cases in the six countries. Overall, scenarios 10 and 11 perform the best for all countries. The relative averted cases (RACs) are computed as the relative difference in the total number of cases (over 2024-2050) under the base scenario and the total number of cases for a certain scenario. These RACs and their interval estimates are depicted in Fig. 8 for the six countries.

**Figure 8.**
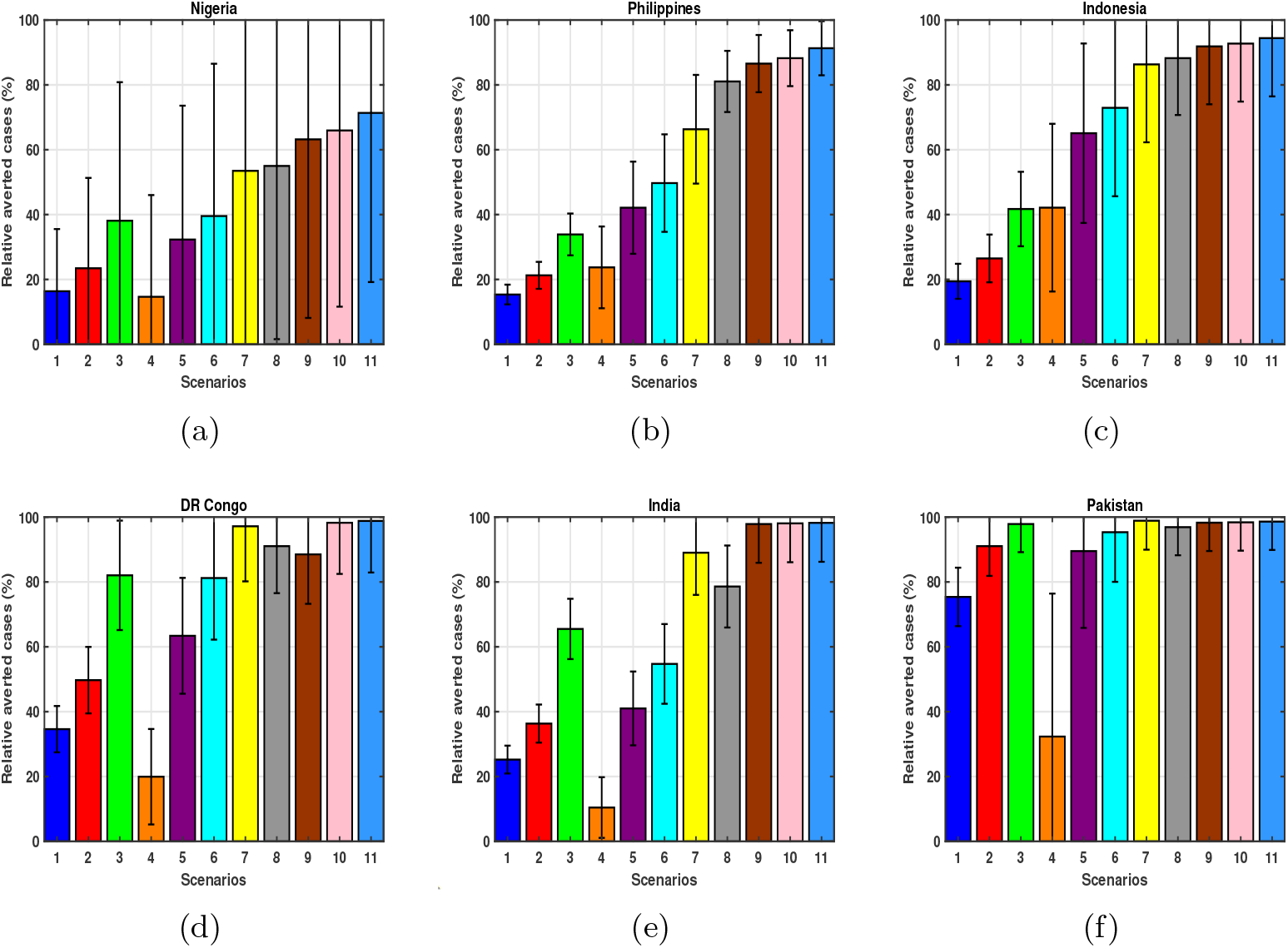
Different color bars represent percent of relative averted cases from 2024 to 2050, for different Scenarios, with respect to the base scenario, as discussed in the text. The averted cases are shown for the countries Nigeria, Philippines, Indonesia, DR Congo, India and Pakistan respectively.

As mentioned above, Nigeria performs the worst among all countries, reporting less than 80% RAC even for scenario 11. Philippines and Indonesia show at least 80% RACs for Scenarios 8-11. DR Congo measles case counts are greatly affected by combining RI with high-frequency SIAs, reaching at least 80% RAC for scenarios 3, 6-11. India shows a reasonable trend of decreasing cases of measles even under the base scenario of RI activities, thus, we note an above 80% RAC value for enhanced scenarios 7-11. Except for scenarios 1 and 4, in Pakistan, all others show more than 80% RAC value.

To better understand the interaction between RACs and year-to-elimination (i.e., absence of endemic measles cases for a period of ≥ 12 months with no further resurgence), we refer to Fig. 9. We observe that scenarios with zero dose vaccination for measles (scenarios 8-11) perform better than other scenarios (RI + SIA, routine MCV1 preponed + MCV2 + SIA) in terms of RACs except in Nigeria. It is also observed that these same scenarios lead to the elimination of measles in all countries except Nigeria. Among the zero-dose scenarios, scenario-11 outperforms other settings. However, scenario 9 is much more economically viable as we are implementing an RI with SIAs every three years and MCV0 for 6-9 months children every three years.

**Figure 9.**
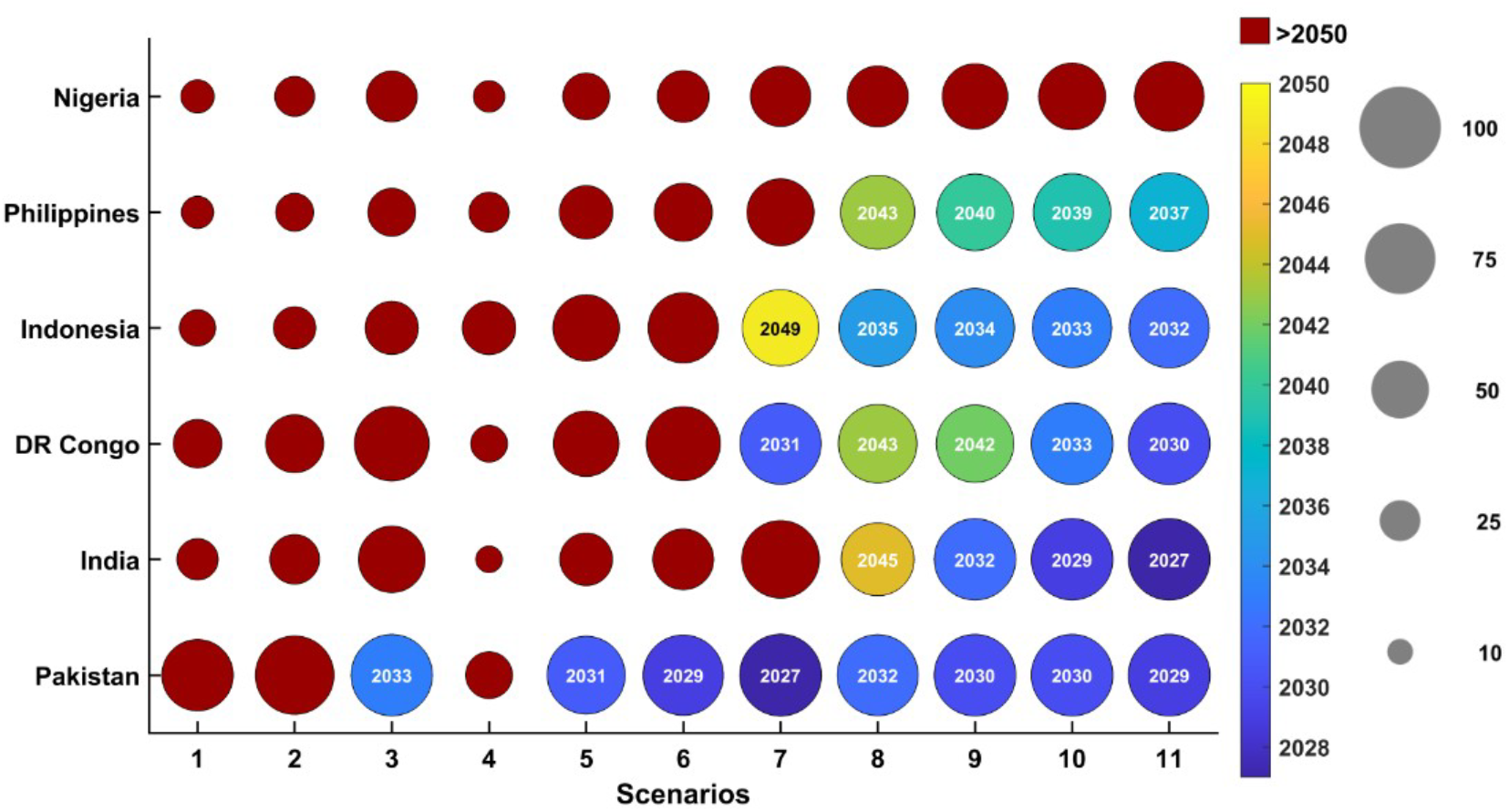
Relative averted cases and year to elimination under different scenarios for the six countries. The size of the bubbles indicates the RACs and the colour of the bubbles represents year to elimination for the corresponding scenarios.

In this manuscript, we consider MCV0 as a pulsed vaccination at 6-9 months, similar to an SIA campaign. In terms of actual life experiences, the pulsed delivery scheme is similar to how health ministries temporarily increase the target age during epidemic outbreak periods. As has been mentioned in the Introduction, countries like Sri Lanka and Colombia have traditionally delivered SIAs for measles with target ages starting at six months to quickly plug the immunity gaps. For actual implementation in Sri Lanka see (Response to measles outbreak in Sri Lanka). This assumption is also motivated by WHO recommendations (WHO Table 3: Recommendations for Interrupted or Delayed Routine Immunization), of administering an additional measles vaccine dose for infants aged 6–9 months in outbreak or other high-risk settings rather than as part of routine immunization.

The choice of introducing an extra dose can also be justified by the number of total doses saved till the elimination of the disease. From Figure 9, we see that the implementation of only SIAs (scenarios 1, 2, and 3) with RI does not eliminate the disease even by 2050. However, if we introduce MCV0 at a small coverage of 28% for all six countries, only for 6-9 months, then elimination is achieved by 2045 in all countries, excluding Nigeria. Thus, an addition of MCV0 in the 6-9month age group helps us to eliminate the disease with a much lower number of total doses.

In terms of measles elimination, Strategy 1 and Strategy 2 appear to be less effective, leading to delays in the achievement of elimination in all countries. In contrast, Strategy 3 performs better in most countries, with the potential to eliminate measles everywhere except in Nigeria. Depending on local epidemiological contexts and resource availability, various alternative scenario projections maybe constructed using the proposed mathematical model.

#### 6.3.1. Additional Scenarios

Further, we also looked at some more additional scenarios where we have considered increase in routine immunization coverage in the forecasting period along with elevated SIA and zero dose coverages for all 6 countries. Also, we have added some scenarios for India, where variable age targeted populations are considered for SIA in the forecasting years. These additional scenarios are described below.

- Scenario 12: 95% MCV1 + 95% MCV2, No SIA,
- Scenario 13: MCV1 + MCV2 at usual coverage (same as last year’s coverage) + SIA with 50% coverage once every three years,
- Scenario 14: 95% MCV1 + 95% MCV2, SIA with 50% coverage once every three years,
- Scenario 15: MCV1 + MCV2 at usual coverage (same as last year’s coverage) + 50% zero-dose coverage, No SIA.

As before we calculated the percentage of relative averted cases of these additional scenarios with respect to the baseline scenario (Scenario-0). The corresponding figure is depicted in Fig. 10(a). Scenario 12 with higher RI in the forecasted years shows improvement over the baseline scenario for all countries. We observe that Scenario 13 gives better results than Scenario-1 for all the countries except Pakistan. This may be due to the fact that Pakistan had 80% SIA coverage in the last year and in Scenario-13 we have considered 50% SIA coverage. Comparing the results of scenario 14 to scenario 1, we see an increase in the relative averted cases. This increase for all 6 countries is due to the high MCV1 and MCV2 coverages (95%) assumed in Scenario 14. We also observe that Scenario 15 shows improvements (higher averted cases) over Scenario 8 due to increased MCV0 coverage for all the countries, where the RI coverages have been kept unchanged.

**Figure 10.**
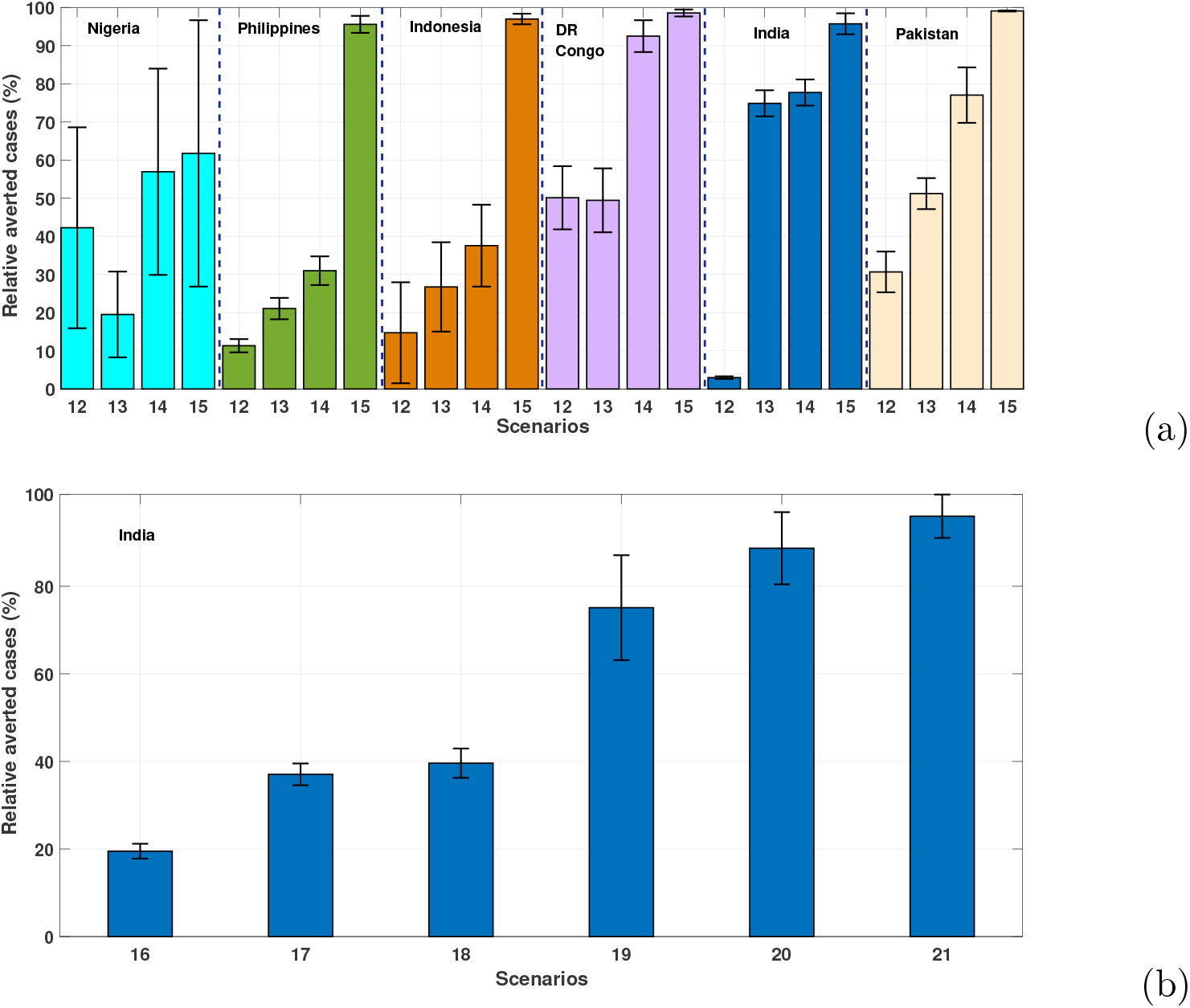
(a) Percentage of relative averted cases from 2024 to 2050, for different additional Scenarios 12-15 in the six countries. (b) Percentage of relative averted cases from 2024 to 2050, for different additional Scenarios 16-21 in India. The averted cases are calculated with respect to the baseline scenario (Scenario-0), as described in Table 3.

Furthermore, we have now looked at different scenarios for India where we have varied the age interval targeted in the SIAs and also the coverage. These additional scenarios are described below. In the additional scenarios (16-21), usual RI coverage is assumed (same as last year’s coverage) along with SIAs at higher coverages and different age groups. The scenarios are as follows:

- Scenario 16 (SIA every three years to the age group 6 months - 10 years with coverage of 16.5%),
- Scenario 17 (SIA every three years to the age group 9 months - 15 years with increased coverage of 33%),
- Scenario 18 (SIA every three years to the age group 6 months - 10 years with increased coverage 33%),
- Scenario 19 (SIA every three years to the age group 9 months - 5 years with increased coverage of 60%),
- Scenario 20 (SIA every three years to the age group 9 months - 10 years with increased coverage of 60%) and
- Scenario 21 (SIA every three years to the age group 6 months - 10 years with increased coverage of 60%).

The corresponding figure is depicted in Fig. 10(b). From scenarios 17 and 18, we observe that SIA in the age group 6 months-10 years averts more cases than in 9 months - 15 years, despite the lower effectiveness of the vaccine in the 6 - 9 months age group. Also, from scenarios 19-21, we see that it is important to cover the 5- 10 years group in the SIA as it leads to higher relative averted cases; this may be due to the case burden in the 5-10 years age group. Within the assumptions of the proposed model, scenarios with 60% SIA coverage among children aged 6 months – 10 years produced the largest projected reductions in measles incidence. These additional scenarios demonstrate the flexibility of the model for analysing a broad range of hypothetical forecasting scenarios.

## 7. Discussion and conclusion

Efforts towards early elimination of measles through various vaccination campaigns are ongoing in all six WHO regions. However, measles remains a major problem for healthcare organizations in many countries with frequent outbreaks. In this study, we formulate an age-stratified measles transmission model to examine the benefits of vaccination to infants younger than 9 months of age. This study investigate the potential epidemiological impact of alternative vaccination scenarios using a mathematical modelling framework.

We establish the positivity and boundedness of the model solutions and analytically obtain the expression of the instantaneous replacement number for the proposed model. From the sensitivity analysis, we observed that an increase in transmission rates significantly increases the total number of measles cases, while an increase in the efficacy of MCV1 coverage significantly decreases total measles cases (Fig. 6). Using reported measles cases, coverages of MCV1, MCV2, SIA and demographic variables from the six countries, we calibrated the proposed model (Fig. 4). The instantaneous replacement number (ℛ_*t*_) is crucial to assess measles transmissibility and evaluate the impact of vaccination strategies [41]. For the six countries, between 2020 and 2023, ℛ_*t*_ values generally ranged from 0.85 to 1.15. The estimated trends of ℛ_*t*_ illustrate how the proposed model can quantify transmission intensity and compare hypothetical intervention scenarios. Furthermore, the modelling frame-work utilized in this study allows for forecasting trends ℛ_*t*_ under various scenarios, offering critical information on the anticipated impact of different vaccination strategies and providing theoretical frameworks to support measles control efforts. In this work, we also analyse the local and global stability of a simplified age-independent version of the original model. Investigating the stability properties of the full-age stratified model remains an immediate goal for our future research.

The scenario analysis for the forecast period (2024–2050) illustrates the ability of the proposed model to compare hypothetical vaccination scenarios under a common modelling framework. Under the model assumptions, scenarios incorporating MCV0 generally resulted in larger projected reductions in measles incidence than those without MCV0. These findings represent comparative model-based results and should not be interpreted as direct recommendations for implementation.

Another important note is that the vaccination frequencies considered in the SIA scenarios are intended to represent population-level coverage and should not be interpreted as implying that the same individual receives a vaccine during every campaign. In practice, repeated vaccination of the same individual in every successive campaign is extremely unlikely. Suppose that, in each annual SIA, a proportion *k*% of the target population is vaccinated, with individuals selected independently from one campaign to the next. Let 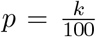, denote the probability that a given individual is selected for vaccination in a single SIA campaign. Then, the probability that the same individual is selected in all *Y* annual campaigns is *p*^*Y*^. Since the annual vaccination coverage satisfies 0 *< p <* 1 in practice, the quantity *p*^*Y*^ decreases exponentially as *Y* increases, making such an extreme vaccination history exceedingly unlikely. Therefore, the proposed model is intended to evaluate the population-level epidemiological impact of repeated SIAs rather than to recommend multiple MCV doses for the same individual. Such extreme individual-level vaccination histories are medically undesirable and lie beyond the scope of the present population-level modelling framework.

A limitation of our study is the assumption of independent coverage for SIAs. In reality, subsequent vaccine doses are often highly dependent on prior dosing, as the same public health system delivers both and may repeatedly miss ‘zero-dose’ children due to systemic barriers. While our model restricts routine MCV2 to those who received MCV1, it distributes SIA doses proportionally across susceptible and previously vaccinated compartments. This assumption of independence might overestimate the efficiency of SIAs in reaching chronically unreached populations. Another limitation of our mathematical formulation is the assumption of independent coverage probabilities for sequential doses. For instance, while the model structurally restricts routine MCV2 to those who previously received MCV1 (the vaccine failure compartment), applying the standard population coverage rate to this group assumes independence. In reality, as demonstrated by Portnoy et al. [42], coverage is highly correlated; an infant who receives an initial dose (e.g., MCV0 or MCV1) has a significantly higher probability of returning for subsequent doses compared to the general population. This assumption of independence may lead the model to underestimate the clustering of immunity in reached populations and overestimate the program’s ability to reach previously missed children. Future modelling efforts should incorporate correlated coverages and heterogeneous access patterns to capture these dynamics more realistically. Future work should also assess the cost-effectiveness of the proposed scenarios using country-specific data.

Finally, this study proposes a mathematical modelling framework for evaluating various hypothetical vaccination scenarios to direct future elimination goals. However, the operational feasibility of any scenario and their practical implementation in real life, should be assessed carefully through collaboration between modellers and policymakers.

## Data Availability

Publicly available data were used, which has been indicated in the manuscript.

## Ethics

This work did not require ethical approval from a human subject or animal welfare committee.

## Data and code availability

The sources of all data used during this study are cited in the article. The codes are available at the following GitHub link: https://github.com/nsamiran/Age-stratified-model.git.

## Declaration of AI use

We have not used AI-assisted technologies in creating this article.

## Disclaimer

The work/opinion is based solely on the research findings of the authors and not the opinion of any government.

## Acknowledgments

The authors are grateful to the learned reviewer for constructive comments and suggestions. Indrajit Ghosh and Siuli Mukhopadhyay were supported by the Gates Foundation (INV-044445).

## Appendix A. Age-independent model with constant parameters

The age-independent version of the model with *κ* = 1 is given by:

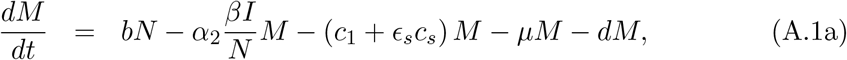

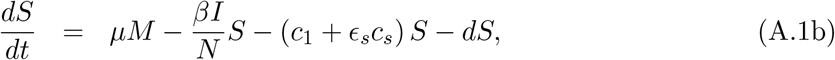

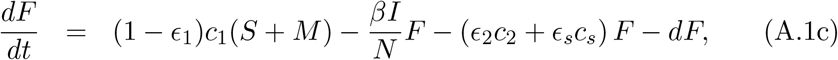

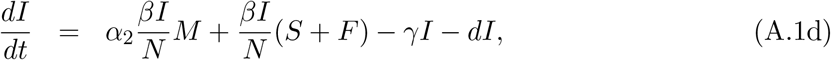

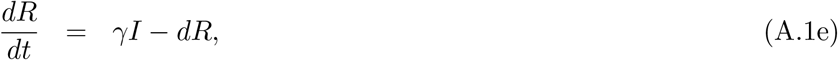

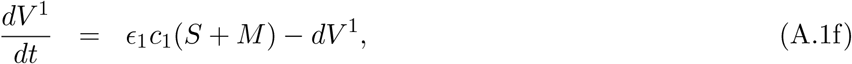

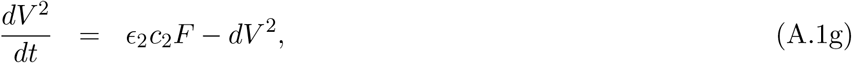

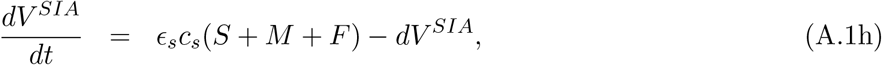

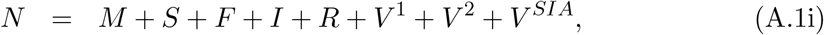

where, all variables and parameters in the age-independent population retain the same meanings as in the age-dependent model (1), where they were previously denoted with the subscript *i* to indicate age group specificity. A flow diagram of the model with and without births and death terms is presented in Fig. A.11. The positivity and boundedness of the system A.1 follow similarly from the proof of positivity and boundedness of the main system (1).

**Figure A.11:**
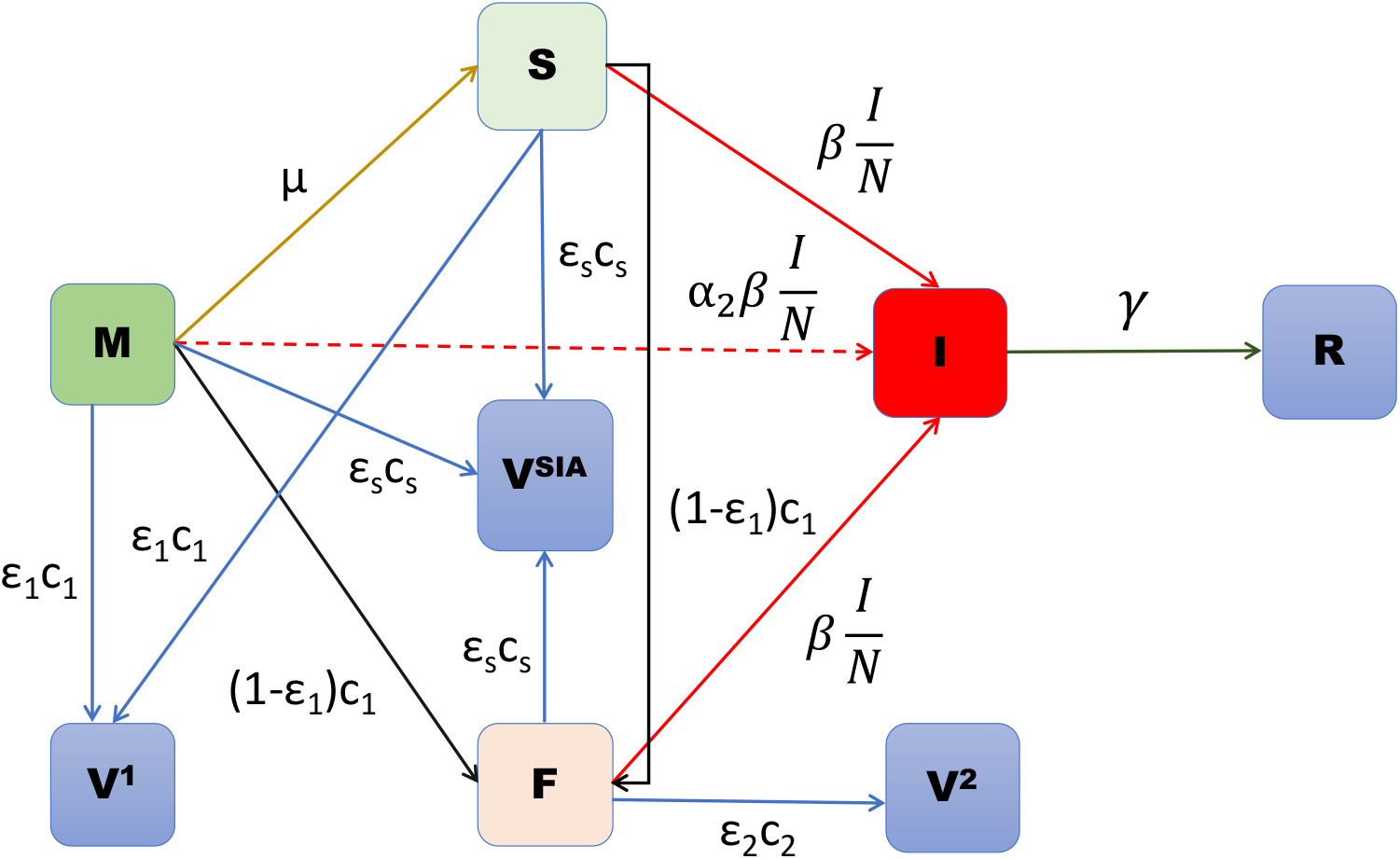
Flow diagram of the age-independent model without births and deaths.

### Appendix A.1. Disease-free equilibrium

If we assume that *b* = *d* then *N* (*t*) = *constant* for all *t* ≥ 0. Under this assumption, the disease-free equilibrium (DFE) point is given by ℰ_∗_ = (*M*_∗_, *S*_∗_, *F*_∗_, *I*_∗_, *R*_∗_, *V* ^1^, *V* ^2^, *V* ^*SIA*^) where

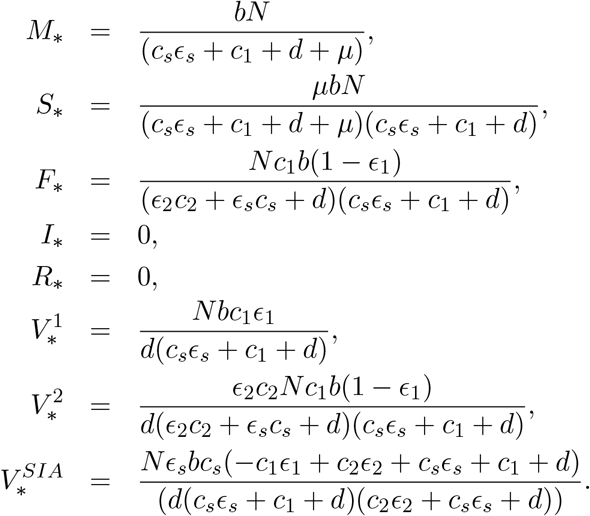

### Appendix A.2. Basic reproduction number

We use the next generation matrix approach [43], to derive the basic reproduction number for the age-independent model (A.1). We consider the compartments *M*, *S, F*, *R, V* ^1^, *V* ^2^, *V* ^*SIA*^ as the non-infected compartments and the compartment *I* as the infected compartment. Then the matrix ℱ corresponding to the new infection and the matrix *V* corresponding to the outflow from the infected compartment are given by:

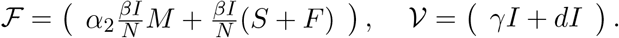

The Jacobian of ℱ and *V* evaluated at the DFE (*M*_∗_, *S*_∗_, *F*_∗_, *I*_∗_, *R*_∗_, *V* ^1^, *V* ^2^, *V* ^*SIA*^) is given by:

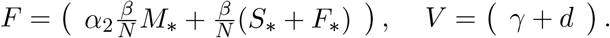

The basic reproduction number is the spectral radius of the matrix *FV* ^−1^ and is given by:

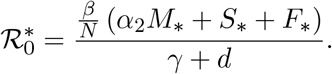

The plot of 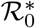 in the parametric space of *c*_1_ and *β/N* is shown in Figure B.18.

### Appendix A.3. Local stability of the DFE

#### Theorem 1.

*The DFE is locally asymptotically stable if* 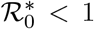 *and unstable if* 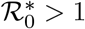.

*Proof*. The Jacobian matrix of the system A.1 around the DFE is given by:

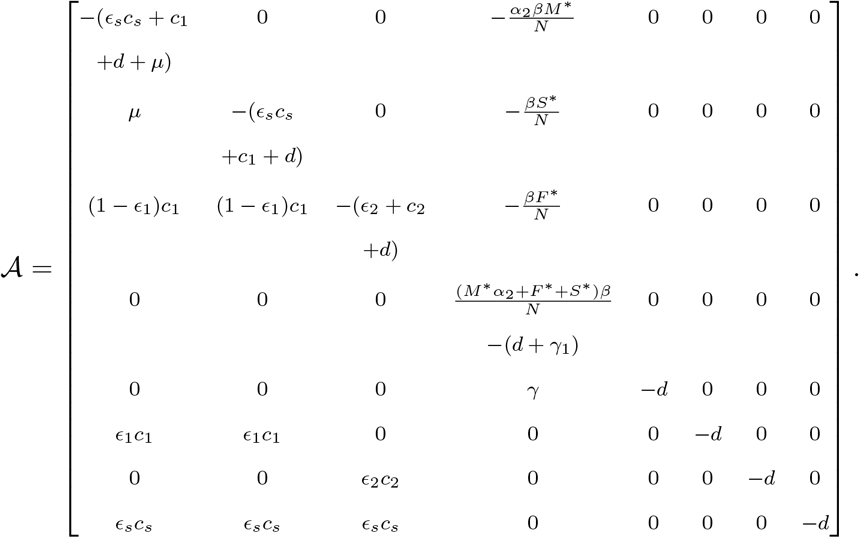

The eigen values of *A* are: −*d*, −*d*, −*d*, −*d* and the rest of the eigen values are determined by the eigen values of the following matrix:

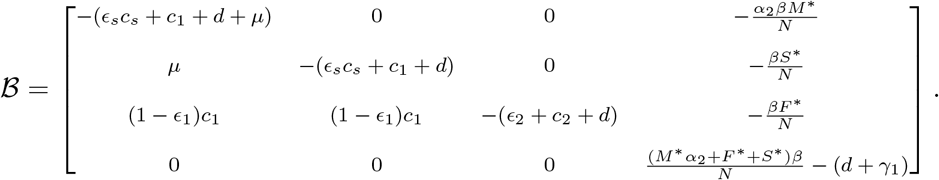

Now the eigen values of *B* are given by:

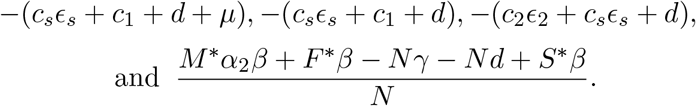

Consequently, all the eigen values of *A* are negative except the last one 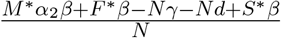 which can be written as 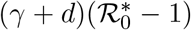. Hence, if 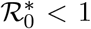 the DFE ℰ_∗_ is locally asymptotically stable and if 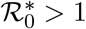 the DFE *E*_∗_ is unstable.

### Appendix A.4. Global stability of the DFE

In this subsection we study the global stability of the disease-free equilibrium. We have the following theorem:

#### Theorem 2.

*The DFE E*_∗_ *is globally asymptotically stable if* 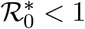.

*Proof*. To prove this theorem we follow the approach described in [44]. We rewrite the system (A.1) in the following form:

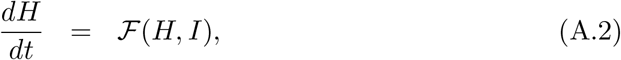

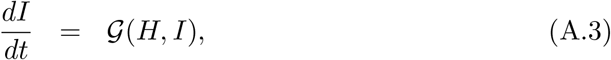

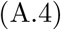

where, 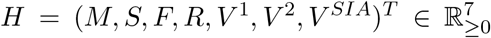 (corresponding to the uninfected compartments), *I* is the infected compartment as defined in the model (A.1) and

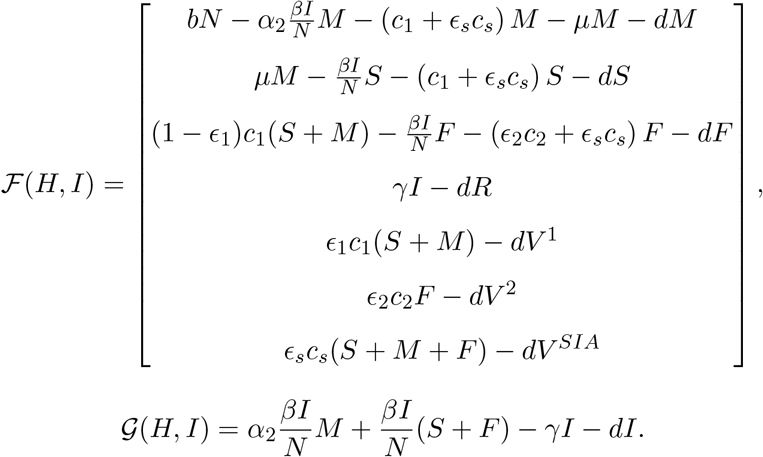

Then the sufficient conditions for the global asymptotic stability of the DFE ℰ_∗_ = (*H*_∗_, 0) are as follows [44]:

1. 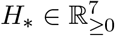 is globally asymptotically stable for the system 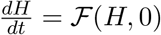.
2. lim_*t*→∞_ *I*(*t*) = 0.

Now,

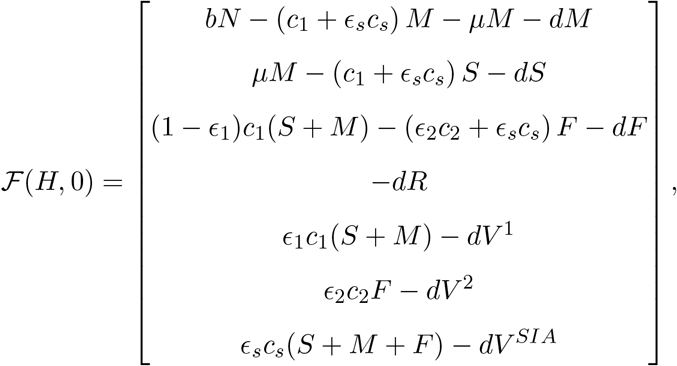

can be written in the form

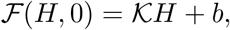

where,

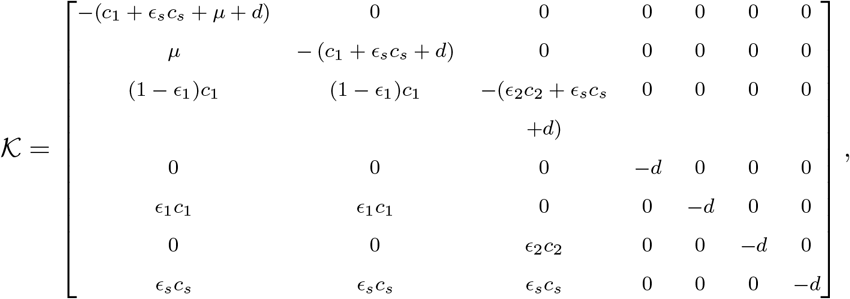

and

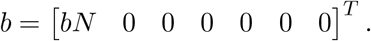

Observe that all the eigen values of *K* are negative, and consequently, *H*_∗_ is globally asymptotically stable for the system 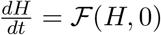.

Now,

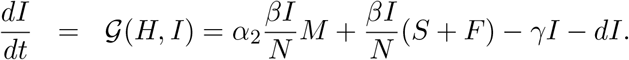

From the first equation of the system (A.1) we get,

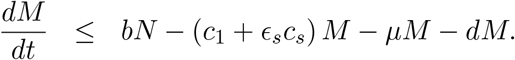

Consider the comparison equation

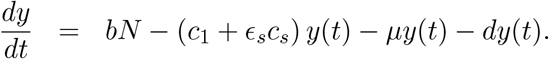

Note that 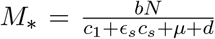 is a globally asymptotically stable equilibrium of the above comparison equation. Hence, for any given *ϵ >* 0 there exists *t*_1_ *>* 0 such that,

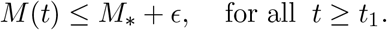

Now from the second equation of the system (A.1), for *t* ≥ *t*_1_, we get,

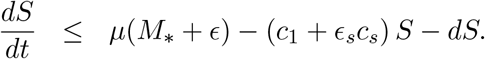

Consider the corresponding comparison equation

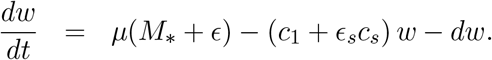

It is easy to observe that

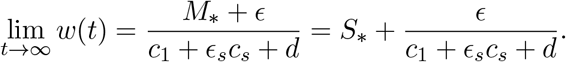

Hence, there exists *t*_2_ *> t*_1_ *>* 0 such that

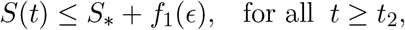

where, 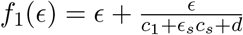.

Now for *t* ≥ *t*_2_, from the third equation of the system (A.1), we get,

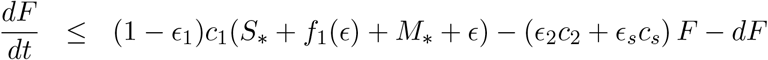

Note that for the corresponding comparison equation

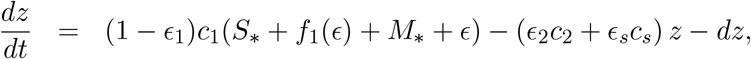

we obtain

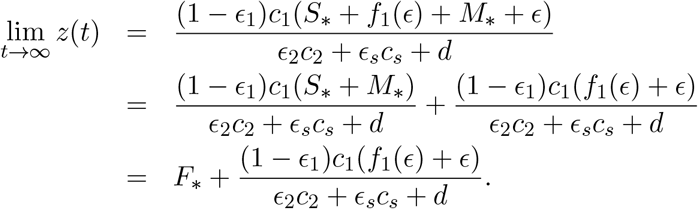

Thus there exists *t*_3_ *> t*_2_ *>* 0 such that

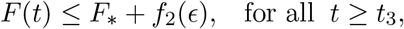

where 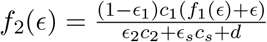.

Now from the equation of *I* in the system (A.1), for *t* ≥ *t*_3_ we get:

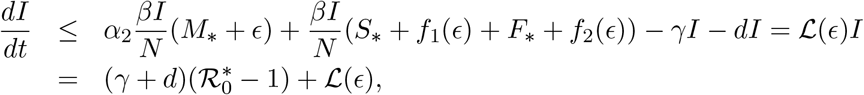

where, 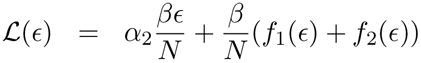.

Since, 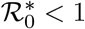 and 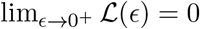. Moreover, note that initial choice of *ϵ >* 0 was arbitrary. Hence, we can choose *ϵ >* 0 in such a way that 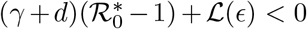. Thus we can conclude that lim_*t*→∞_ *I*(*t*) = 0. This completes the proof.

### Appendix A.5. Existence of endemic equilibrium

Suppose the endemic equilibrium point is denoted by 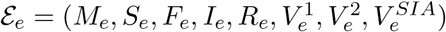. Then we have,

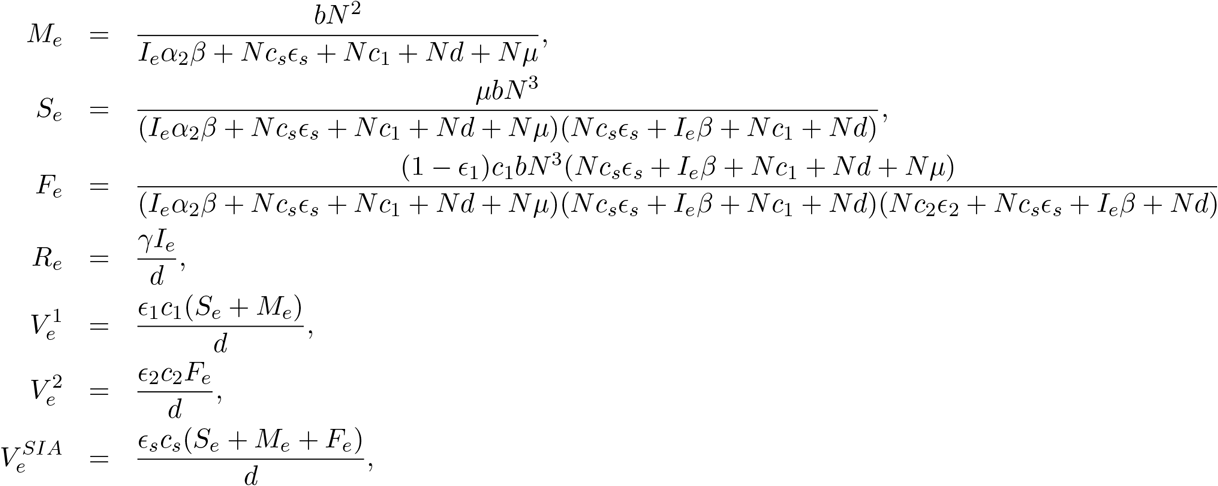

and the solution of *I*_*e*_ is obtained by the following equation:

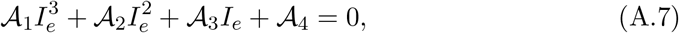

where

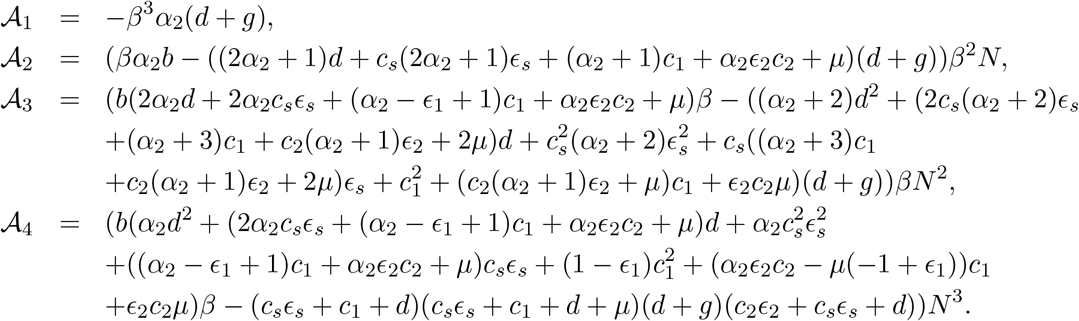

Note that if the equation (A.7) has positive solution then there exists positive endemic equilibrium 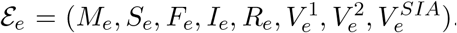. Due to the complexity of the equation (A.7), we are unable to perform further analytical study. However, using equivalent parameter setup as in Table 1, numerically we studied the existence of forward bifurcation as shown in Figure (B.19).

## Appendix B. Supplementary figures

## Appendix C. Supplementary tables

**Table. C4:**
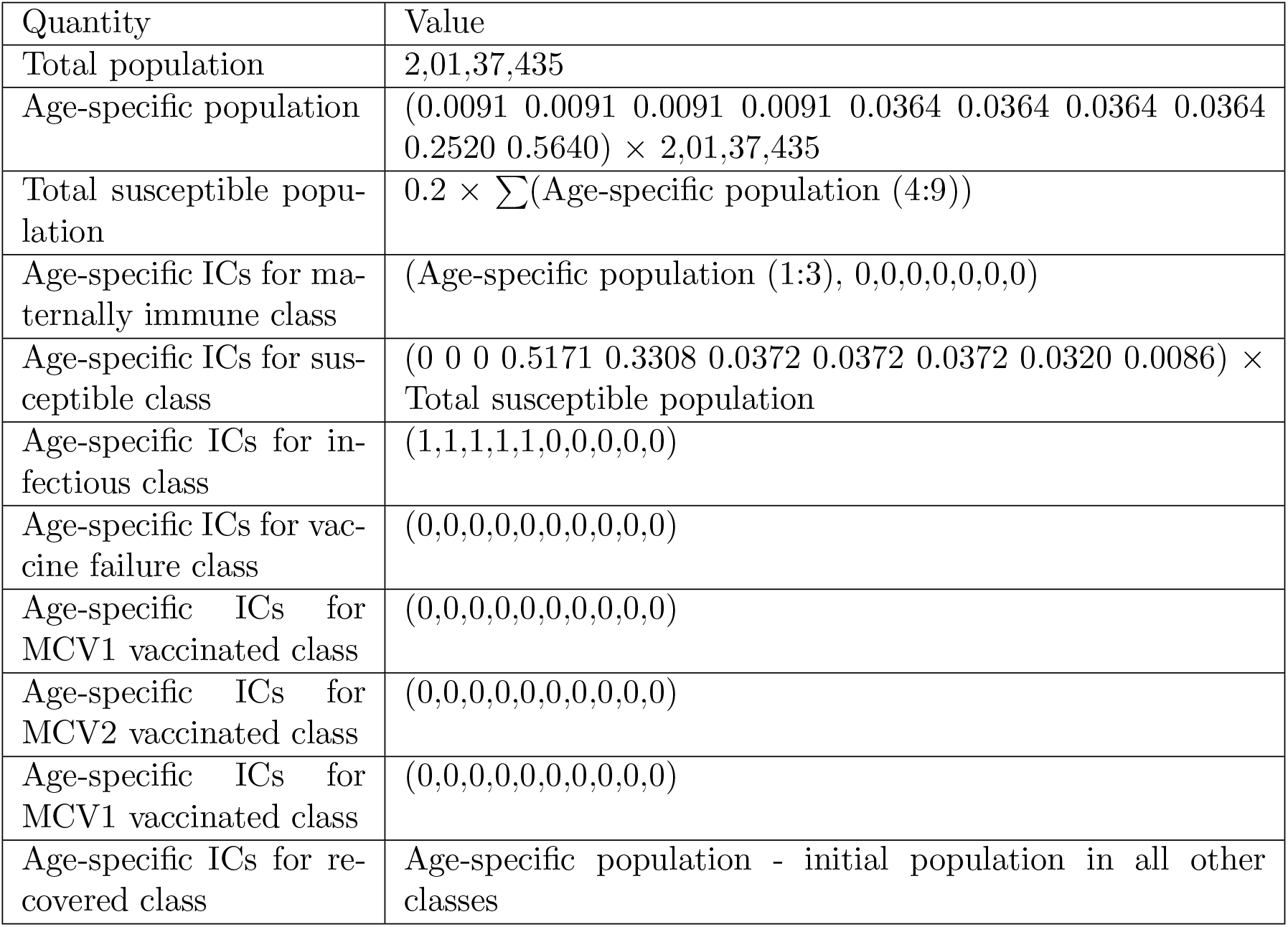
Initial population, age-specific population and initial population sizes of the compartments for DR Congo in the year 1970. Total population and age-specific populations are obtained from https://www.populationpyramid.net/democratic-republic-of-the-congo/1970/. Proportions for age-specific susceptibles are assumed to be the proportion of the infection in the age classes, except for the first three age groups (https://www.healthdata.org/research-analysis/gbd-publications).

| Quantity | Value |
| --- | --- |
| Total population | 2,01,37,435 |
| Age-specific population | $(0.0091 \ 0.0091 \ 0.0091 \ 0.0091 \ 0.0364 \ 0.0364 \ 0.0364 \ 0.0364 \ 0.2520 \ 0.5640) \times 2,01,37,435$ |
| Total susceptible population | $0.2 \times \sum(\text{Age-specific population (4:9)})$ |
| Age-specific ICs for maternally immune class | $(\text{Age-specific population (1:3)}, 0,0,0,0,0,0)$ |
| Age-specific ICs for susceptible class | $(0 \ 0 \ 0 \ 0.5171 \ 0.3308 \ 0.0372 \ 0.0372 \ 0.0372 \ 0.0320 \ 0.0086) \times \text{Total susceptible population}$ |
| Age-specific ICs for infectious class | $(1,1,1,1,1,0,0,0,0,0)$ |
| Age-specific ICs for vaccine failure class | $(0,0,0,0,0,0,0,0,0,0)$ |
| Age-specific ICs for MCV1 vaccinated class | $(0,0,0,0,0,0,0,0,0,0)$ |
| Age-specific ICs for MCV2 vaccinated class | $(0,0,0,0,0,0,0,0,0,0)$ |
| Age-specific ICs for MCV1 vaccinated class | $(0,0,0,0,0,0,0,0,0,0)$ |
| Age-specific ICs for recovered class | Age-specific population - initial population in all other classes |

**Table. C5:** Initial estimated parameters for the six countries.

| Parameters<br>Countries | $\alpha_1^1$ | $\alpha_1^2$ | $\alpha_1^3$ | $\alpha_1^4$ | $\kappa$ | $\rho$ |
| --- | --- | --- | --- | --- | --- | --- |
| Nigeria | 1.1639 | 0.011 | 0.25 | 0.001 | 1 | 0.0244 |
| Philippines | 1.3 | 0.09 | 0.08 | 0.008 | 0.99 | 0.009 |
| Indonesia | 1.2514 | 0.09 | 0.08 | 0.0008 | 1 | 0.009 |
| DR Congo | 1.0274 | 0.2818 | 0.0984 | 0.0265 | 0.9093 | 0.0025 |
| India | 0.8249 | 0.5971 | 0.0658 | 0.0012 | 0.9 | 0.0045 |
| Pakistan | 1.6324 | 0.0904 | 0.08 | 0.0008 | 0.5904 | 0.009 |

**Figure B.12:**
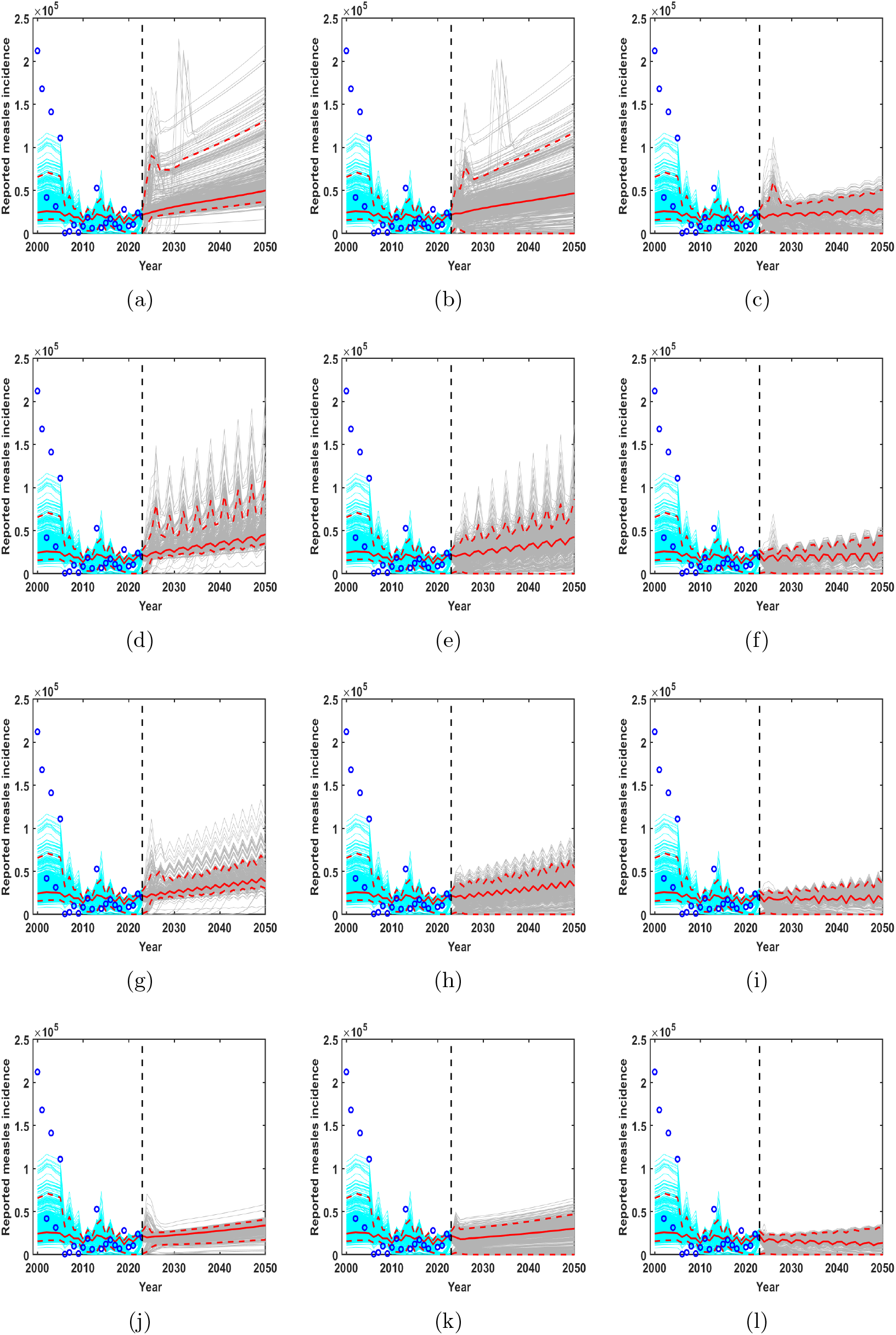
Nigeria. Model fitting for the time period 2000 to 2023 and forecasting for different scenarios from 2024 to 2050 are shown with 95% confidence intervals. Panels (a), (d), (g), and (j) correspond to the base scenario and Scenarios 1, 2, and 3, respectively; (b), (e), (h), and (k) correspond to Scenarios 4, 5, 6, and 7, respectively; and (c), (f), (i), and (l) correspond to Scenarios 8, 9, 10, and 11, respectively.

**Figure B.13:**
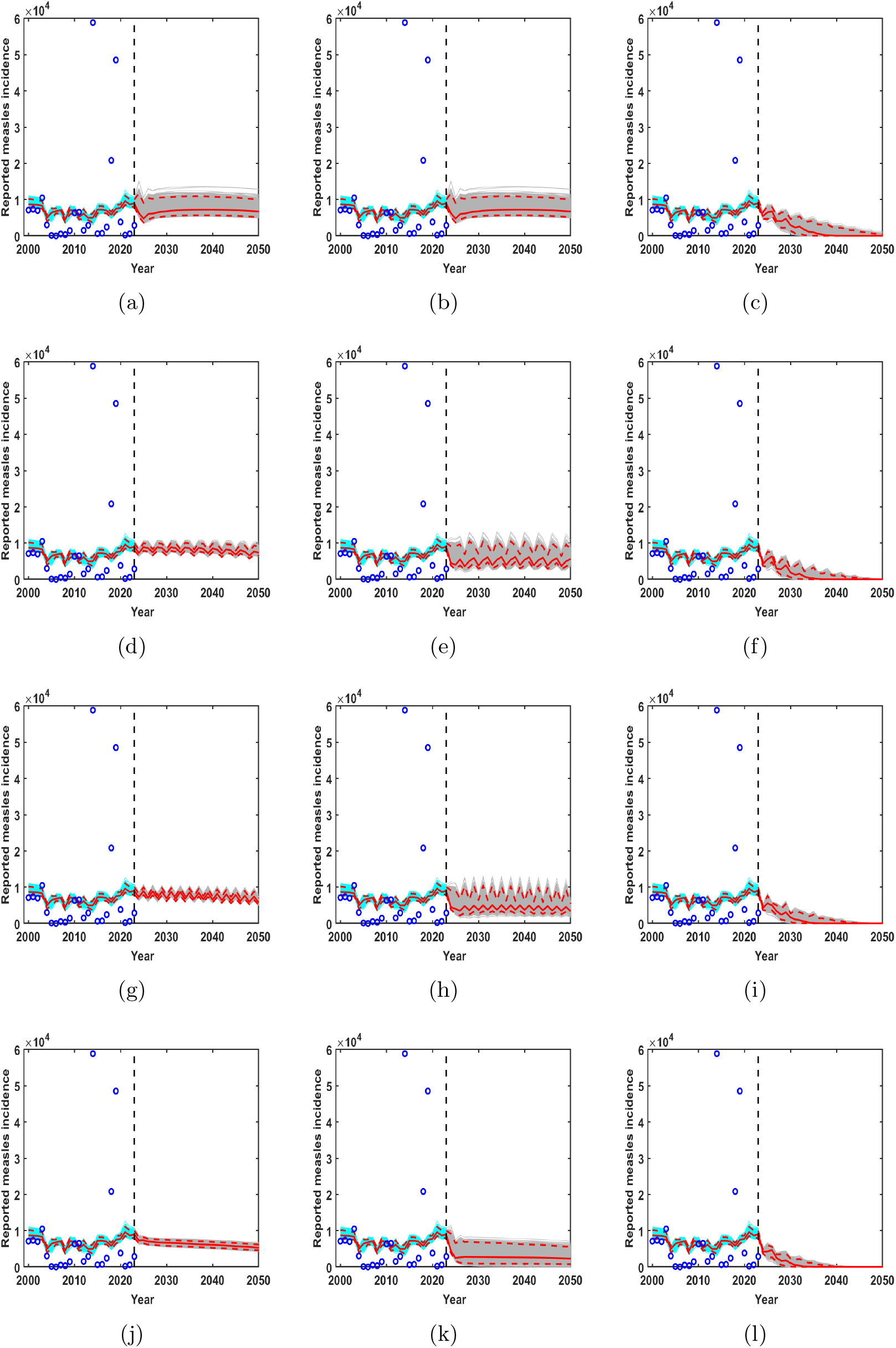
Philippines. Model fitting for the time period 2000 to 2023 and forecasting for different scenarios from 2024 to 2050 are shown with 95% confidence intervals. Panels (a), (d), (g), and (j) correspond to the base scenario and Scenarios 1, 2, and 3, respectively; (b), (e), (h), and (k) correspond to Scenarios 4, 5, 6, and 7, respectively; and (c), (f), (i), and (l) correspond to Scenarios 8, 9, 10, and 11, respectively.

**Figure B.14:**
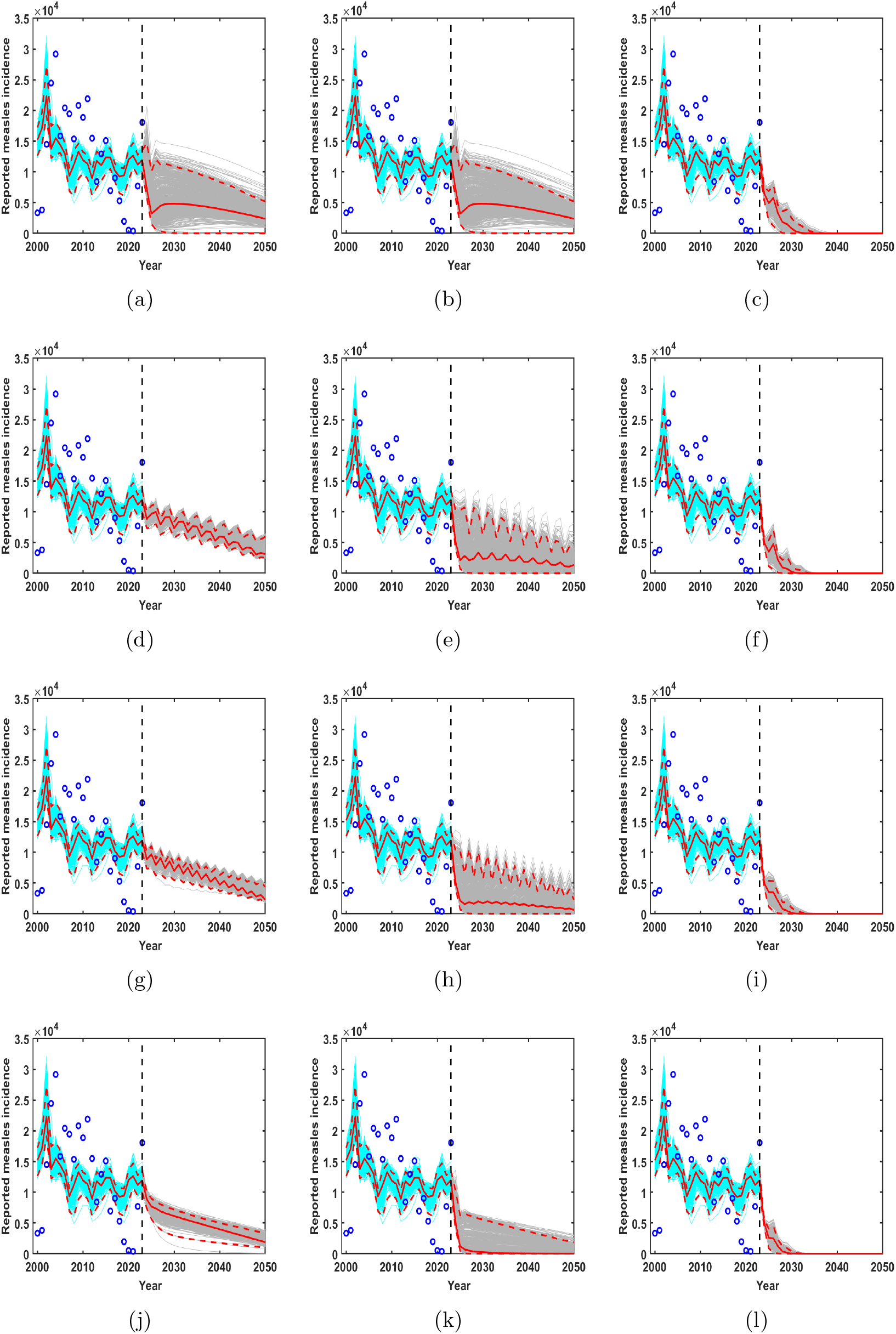
Indonesia. Model fitting for the time period 2000 to 2023 and forecasting for different scenarios from 2024 to 2050 are shown with 95% confidence intervals. Panels (a), (d), (g), and (j) correspond to the base scenario and Scenarios 1, 2, and 3, respectively; (b), (e), (h), and (k) correspond to Scenarios 4, 5, 6, and 7, respectively; and (c), (f), (i), and (l) correspond to Scenarios 8, 9, 10, and 11, respectively.

**Figure B.15:**
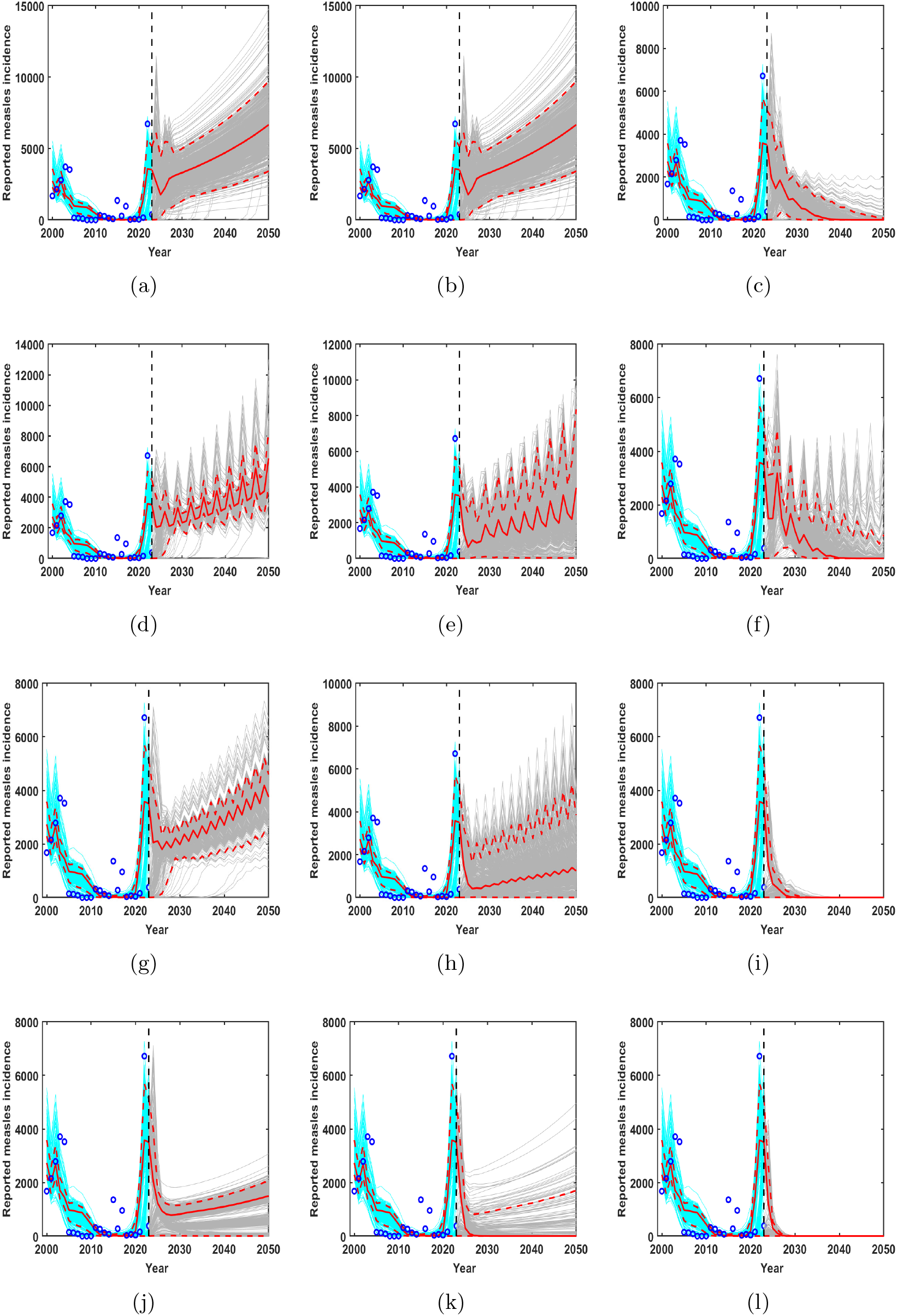
DR Congo. Model fitting for the time period 2000 to 2023 and forecasting for different scenarios from 2024 to 2050 are shown with 95% confidence intervals. Panels (a), (d), (g), and (j) correspond to the base scenario and Scenarios 1, 2, and 3, respectively; (b), (e), (h), and (k) correspond to Scenarios 4, 5, 6, and 7, respectively; and (c), (f), (i), and (l) correspond to Scenarios 8, 9, 10, and 11, respectively.

**Figure B.16:**
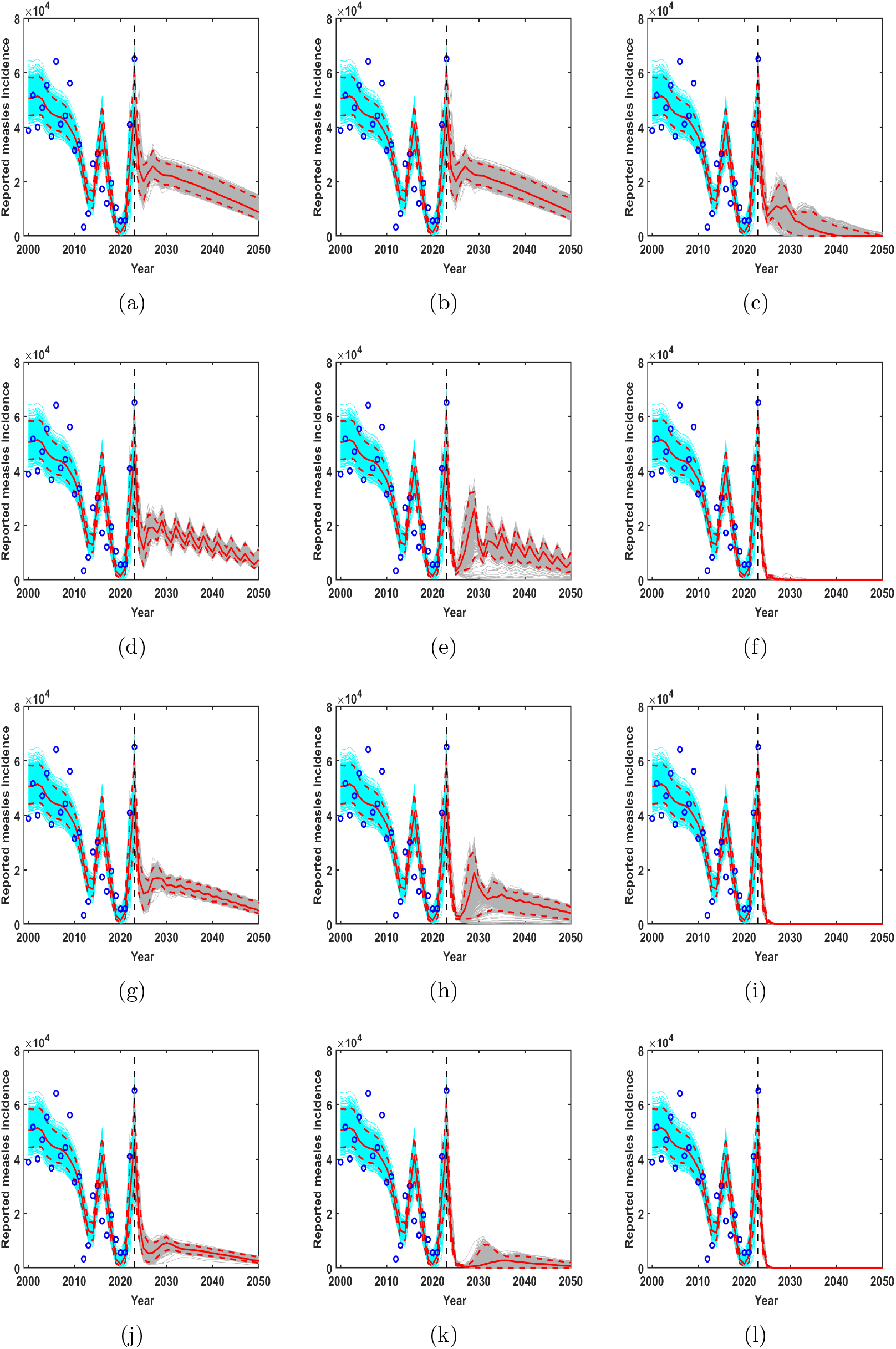
India. Model fitting for the time period 2000 to 2023 and forecasting for different scenarios from 2024 to 2050 are shown with 95% confidence intervals. Panels (a), (d), (g), and (j) correspond to the base scenario and Scenarios 1, 2, and 3, respectively; (b), (e), (h), and (k) correspond to Scenarios 4, 5, 6, and 7, respectively; and (c), (f), (i), and (l) correspond to Scenarios 8, 9, 10, and 11, respectively.

**Figure B.17:**
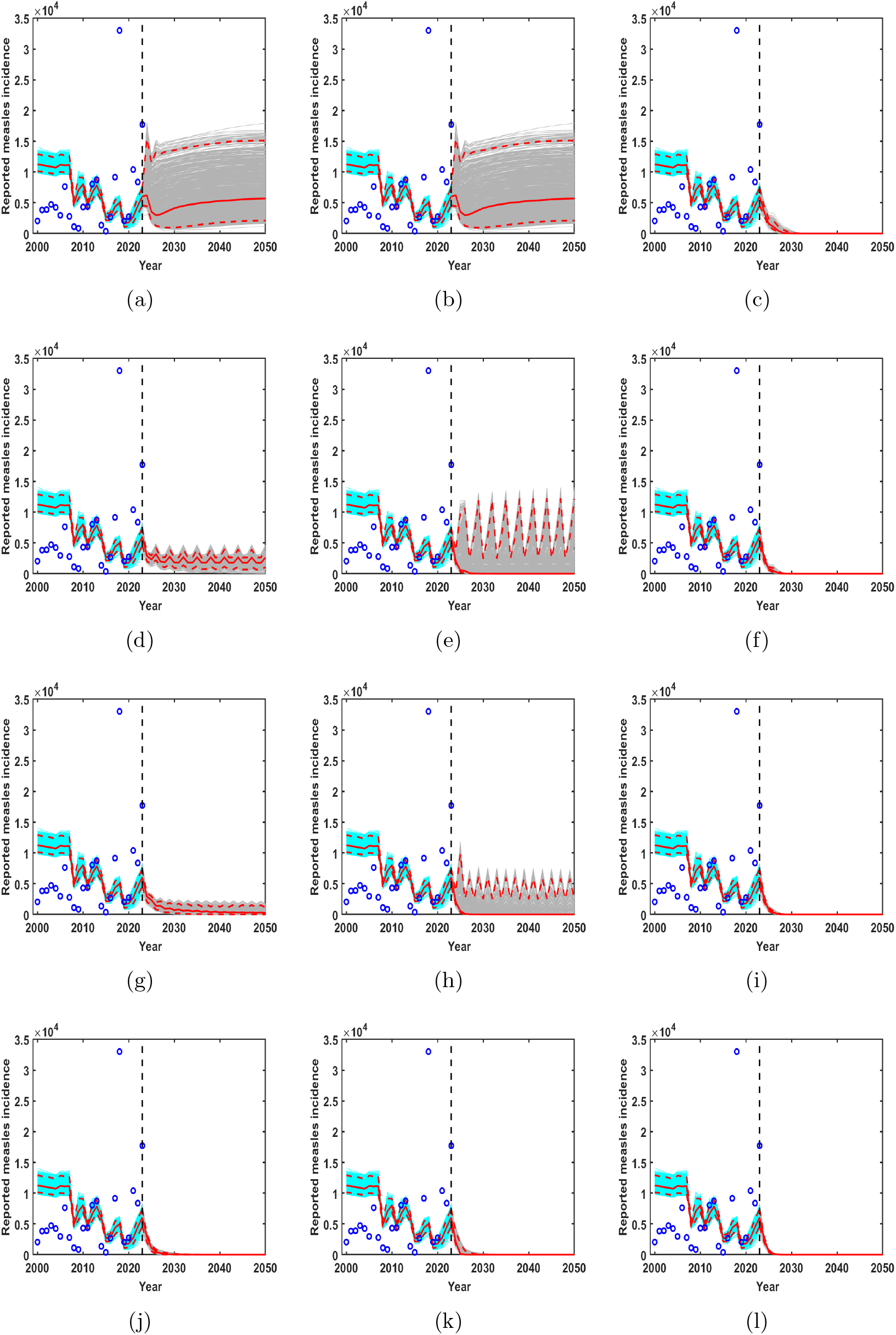
Pakistan. Model fitting for the time period 2000 to 2023 and forecasting for different scenarios from 2024 to 2050 are shown with 95% confidence intervals. Panels (a), (d), (g), and (j) correspond to the base scenario and Scenarios 1, 2, and 3, respectively; (b), (e), (h), and (k) correspond to Scenarios 4, 5, 6, and 7, respectively; and (c), (f), (i), and (l) correspond to Scenarios 8, 9, 10, and 11, respectively.

**Figure B.18:**
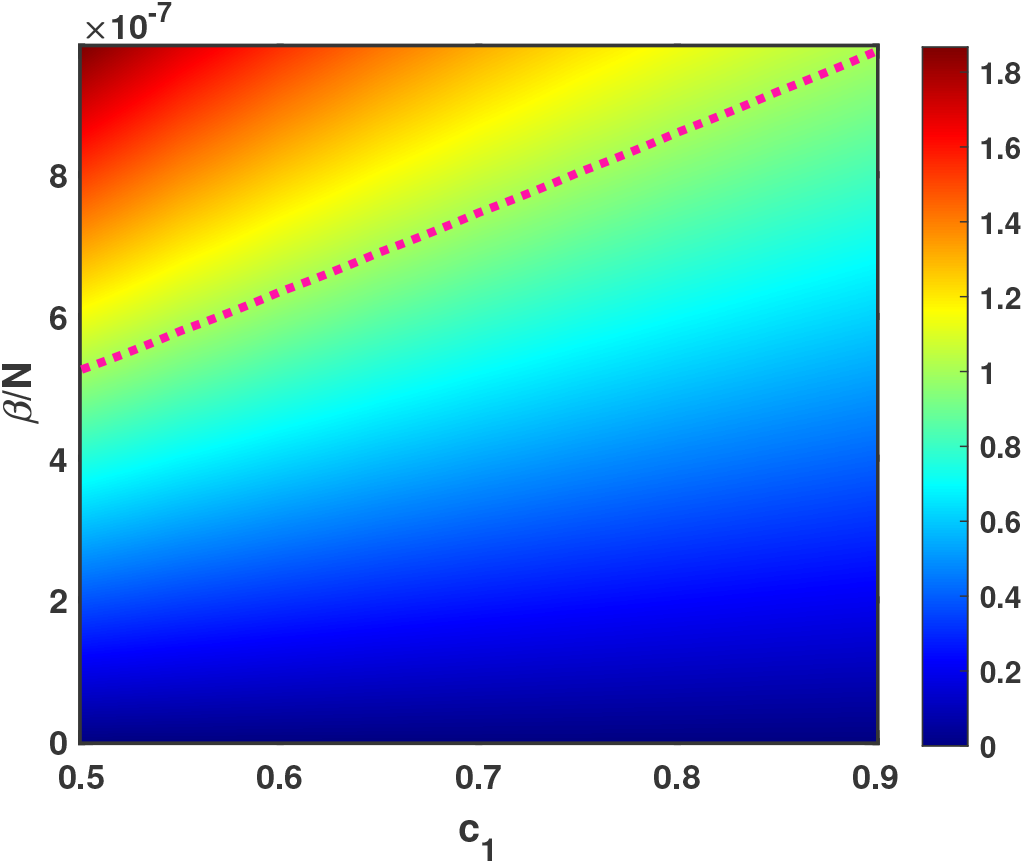
Plot of 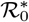 in the parameter space. The other parameters are chosen equivalently as in Table 1.

**Figure B.19:**
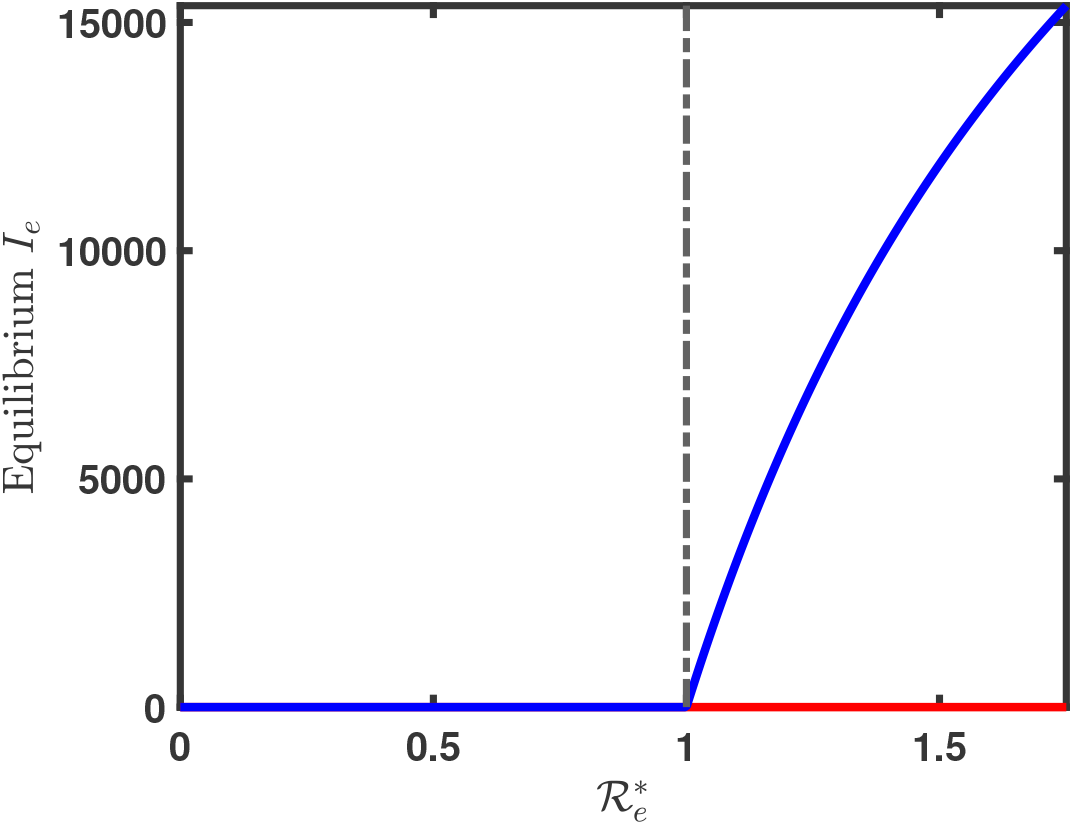
Forward bifurcation diagram. Blue color corresponds to stable equilibrium point and red color corresponds to unstable equilibrium point.

## References

[1] Abhishek Pandey and Alison P Galvani. Exacerbation of measles mortality by vaccine hesitancy worldwide. The Lancet Global Health, 11(4):e478–e479, 2023.

[2] Megan Auzenbergs, Han Fu, Kaja Abbas, Simon R Procter, Felicity T Cutts, and Mark Jit. Health effects of routine measles vaccination and supplementary immunisation activities in 14 high-burden countries: a dynamic measles immunization calculation engine (dynamice) modelling study. The Lancet Global Health, 11(8):e1194–e1204, 2023.

[3] Wei Wang, Megan O’Driscoll, Qianli Wang, Sihong Zhao, Henrik Salje, and Hongjie Yu. Dynamics of measles immunity from birth and following vaccination. Nature Microbiology, pages 1–10, 2024.

[4] Laura M Nic Lochlainn, Brechje de Gier, Nicoline van der Maas, Rob van Bin-nendijk, Peter M Strebel, Tracey Goodman, Hester E de Melker, William J Moss, and Susan JM Hahné. Effect of measles vaccination in infants younger than 9 months on the immune response to subsequent measles vaccine doses: a systematic review and meta-analysis. The Lancet Infectious Diseases, 19(11):1246–1254, 2019.

[5] Minal K Patel and Walter A Orenstein. Classification of global measles cases in 2013–17 as due to policy or vaccination failure: a retrospective review of global surveillance data. The Lancet Global Health, 7(3):e313–e320, 2019.

[6] Moss WJ Measles. Lancet (london england). Google Scholars, 390(10111):2490–502, 2017.

[7] WHO. Epidemiology of measles in infants younger than 6 months: analysis of surveillance data 2011-2016 an analysis of the epidemiology of measles in infants younger than 6 months was conducted by the U.S. CDC and WHO using case-based measles surveillance. 2017. Available at: https://api.semanticscholar.org/CorpusID:51938638.

[8] E Leuridan, M Sabbe, and P Van Damme. Measles outbreak in europe: susceptibility of infants too young to be immunized. Vaccine, 30(41):5905–5913, 2012.

[9] Elke Leuridan, Niel Hens, V Hutse, M Ieven, Marc Aerts, and P Van Damme. Early waning of maternal measles antibodies in era of measles elimination: longitudinal study. Bmj, 340, 2010.

[10] A Gagneur and D Pinquier. Spotlight on measles 2010: Timely administration of the first dose of measles vaccine in the context of an ongoing measles outbreak in france. Eurosurveillance, 15(41):19689, 2010.

[11] Vaccination schedule for measles. https://immunizationdata.who.int/global/wiise-detail-page/vaccination-schedule-for-measles?ISO_3_CODE=&TARGETPOP_GENERAL=.

[12] Mmr supplementary immunization activity. https://www.epid.gov.lk/responding-to-the-current-measles-outbreak.

[13] T Jacob John and Valsan P Verghese. Time to re-think measles vaccination schedule in india. Indian Journal of Medical Research, 134(3):256–259, 2011.

[14] Laura M Nic Lochlainn, Brechje de Gier, Nicoline van der Maas, Peter M Strebel, Tracey Goodman, Rob S van Binnendijk, Hester E de Melker, and Susan JM Hahné. Immunogenicity, effectiveness, and safety of measles vaccination in infants younger than 9 months: a systematic review and meta-analysis. The Lancet Infectious Diseases, 19(11):1235–1245, 2019.

[15] Matt J Keeling and Pejman Rohani. Modeling infectious diseases in humans and animals. Princeton university press, 2008.

[16] Filippo Trentini, Piero Poletti, Stefano Merler, and Alessia Melegaro. Measles immunity gaps and the progress towards elimination: a multi-country modelling analysis. The Lancet Infectious Diseases, 17(10):1089–1097, 2017.

[17] Stéphane Verguet, Mira Johri, Shaun K Morris, Cindy L Gauvreau, Prabhat Jha, and Mark Jit. Controlling measles using supplemental immunization activities: a mathematical model to inform optimal policy. Vaccine, 33(10):1291–1296, 2015.

[18] Joaquín M Prada, C Jessica E Metcalf, Saki Takahashi, J Lessler, AJ Tatem, and M Ferrari. Demographics, epidemiology and the impact of vaccination campaigns in a measles-free world–can elimination be maintained? Vaccine, 35(11):1488–1493, 2017.

[19] Filippo Trentini, Piero Poletti, Alessia Melegaro, and Stefano Merler. The introduction of ‘no jab, no school’policy and the refinement of measles immunisation strategies in high-income countries. BMC medicine, 17:1–8, 2019.

[20] Han Fu, Kaja Abbas, Petra Klepac, Kevin van Zandvoort, Hira Tanvir, Allison Portnoy, and Mark Jit. Effect of evidence updates on key determinants of measles vaccination impact: a dynamice modelling study in ten high-burden countries. BMC medicine, 19:1–15, 2021.

[21] Amy K Winter, Brian Lambert, Daniel Klein, Petra Klepac, Timos Papadopoulos, Shaun Truelove, Colleen Burgess, Heather Santos, Jennifer K Knapp, Su-san E Reef, et al. Feasibility of measles and rubella vaccination programmes for disease elimination: a modelling study. The Lancet Global Health, 10(10):e1412– e1422, 2022.

[22] Elizabeth Goult, Laura Andrea Barrero Guevara, Michael Briga, and Matthieu Domenech de Cellès. Estimating the optimal age for infant measles vaccination. Nature Communications, 15(1):9919, 2024.

[23] World Health Organization. Measles vaccines: Who position paper – april 2017. weekly epidemiological record [internet]. 92–92(17):205–28, 2017.

[24] Jöel Mossong and Claude P Muller. Modelling measles re-emergence as a result of waning of immunity in vaccinated populations. Vaccine, 21(31):4597–4603, 2003.

[25] FMG Magpantay, AA King, and P Rohani. Age-structure and transient dynamics in epidemiological systems. Journal of the Royal Society Interface, 16(156):20190151, 2019.

[26] Joseph L Mathew, Abram L Wagner, Radha Kanta Ratho, Pooja N Patel, Vanita Suri, Bhavneet Bharti, Bradley F Carlson, Sourabh Dutta, Mini P Singh, and Matthew L Boulton. Maternally transmitted anti-measles antibodies, and susceptibility to disease among infants in chandigarh, india: A prospective birth cohort study. Plos one, 18(10):e0287110, 2023.

[27] Manoj V Murhekar, Nivedita Gupta, Alvira Z Hasan, Muthusamy Santhosh Kumar, V Saravana Kumar, Christine Prosperi, Gajanan N Sapkal, Jeromie Wesley Vivian Thangaraj, Ojas Kaduskar, Vaishali Bhatt, et al. Evaluating the effect of measles and rubella mass vaccination campaigns on seroprevalence in india: a before-and-after cross-sectional household serosurvey in four districts, 2018–2020. The Lancet Global Health, 10(11):e1655–e1664, 2022.

[28] Xia Yang, Lansun Chen, and Jufang Chen. Permanence and positive periodic solution for the single-species nonautonomous delay diffusive models. Computers & mathematics with applications, 32(4):109–116, 1996.

[29] Herbert W Hethcote. The mathematics of infectious diseases. SIAM review, 42(4):599–653, 2000.

[30] Donald W Marquardt. An algorithm for least-squares estimation of nonlinear parameters. Journal of the society for Industrial and Applied Mathematics, 11(2):431–441, 1963.

[31] Thomas F Coleman and Yuying Li. An interior trust region approach for nonlinear minimization subject to bounds. SIAM Journal on optimization, 6(2):418–445, 1996.

[32] Gerardo Chowell. Fitting dynamic models to epidemic outbreaks with quantified uncertainty: A primer for parameter uncertainty, identifiability, and forecasts. Infectious Disease Modelling, 2(3):379–398, 2017.

[33] Kimberlyn Roosa and Gerardo Chowell. Assessing parameter identifiability in compartmental dynamic models using a computational approach: application to infectious disease transmission models. Theoretical Biology and Medical Modelling, 16:1–15, 2019.

[34] Biplab Maity, Bapi Saha, Indrajit Ghosh, and Joydev Chattopadhyay. Model-based estimation of expected time to cholera extinction in lusaka, zambia. Bulletin of Mathematical Biology, 85(7):55, 2023.

[35] Simeone Marino, Ian B Hogue, Christian J Ray, and Denise E Kirschner. A methodology for performing global uncertainty and sensitivity analysis in systems biology. Journal of theoretical biology, 254(1):178–196, 2008.

[36] Thi Huyen Trang Nguyen, Thuong Vu Nguyen, Quang Chan Luong, Thang Vinh Ho, Christel Faes, and Niel Hens. Understanding the transmission dynamics of a large-scale measles outbreak in southern vietnam. International Journal of Infectious Diseases, 122:1009–1017, 2022.

[37] DO Simba and GI Msamanga. Measles vaccine effectiveness under field conditions. a case control study in tabora region, tanzania. Tropical and geographical medicine, 47(5):197–199, 1995.

[38] Who disease outbreak news; measles–indonesia. Available at https://www.who.int/emergencies/disease-outbreak-news/item/2023-DON462.

[39] Fleurette M Domai, Kristal An Agrupis, Su Myat Han, Ana Ria Sayo, Janine S Ramirez, Raphael Nepomuceno, Shuichi Suzuki, Annavi Marie G Villanueva, Eumelia P Salva, Jose Benito Villarama, et al. Measles outbreak in the philip-pines: epidemiological and clinical characteristics of hospitalized children, 2016-2019. The Lancet Regional Health–Western Pacific, 19, 2022.

[40] Niket Thakkar, Syed Saqlain Ahmad Gilani, Quamrul Hasan, and Kevin A Mc-Carthy. Decreasing measles burden by optimizing campaign timing. Proceedings of the National Academy of Sciences, 116(22):11069–11073, 2019.

[41] Fiona M Guerra, Shelly Bolotin, Gillian Lim, Jane Heffernan, Shelley L Deeks, Ye Li, and Natasha S Crowcroft. The basic reproduction number (r0) of measles: a systematic review. The Lancet Infectious Diseases, 17(12):e420–e428, 2017.

[42] Allison Portnoy, Mark Jit, Stéphane Helleringer, and Stéphane Verguet. Impact of measles supplementary immunization activities on reaching children missed by routine programs. Vaccine, 36(1):170–178, 2018.

[43] Pauline Van den Driessche and James Watmough. Reproduction numbers and sub-threshold endemic equilibria for compartmental models of disease transmission. Mathematical biosciences, 180(1-2):29–48, 2002.

[44] Carlos Castillo-Chavez, Zhilan Feng, and Wenzhang Huang. On the computation of r0 and its role on global stability. Mathematical approaches for emerging and reemerging infectious diseases: an introduction, 1:229, 2002.

